# Data-driven multiscale dynamical framework to control a pandemic evolution with non-pharmaceutical interventions

**DOI:** 10.1101/2021.07.28.21260870

**Authors:** Jürgen Reingruber, Andrea Papale, Stéphane Ruckly, Jean-Francois Timsit, David Holcman

## Abstract

Before the availability of vaccines, many countries have resorted multiple times to drastic social restrictions to prevent saturation of their health care system, and to regain control over an otherwise exponentially increasing COVID-19 pandemic. With the advent of data-sharing, computational approaches are key to efficiently control a pandemic with non-pharmaceutical interventions (NPIs). Here we develop a data-driven computational framework based on a time discrete and age-stratified compartmental model to control a pandemic evolution inside and outside hospitals in a constantly changing environment with NPIs. Besides the calendrical time, we introduce a second time-scale for the infection history, which allows for non-exponential transition probabilities. We develop inference methods and feedback procedures to successively recalibrate model parameters as new data becomes available. As a showcase, we calibrate the framework to study the pandemic evolution inside and outside hospitals in France until February 2021. We combine national hospitalization statistics from governmental websites with clinical data from a single hospital to calibrate hospitalization parameters. We infer changes in social contact matrices as a function of NPIs from positive testing and new hospitalization data. We use simulations to infer hidden pandemic properties such as the fraction of infected population, the hospitalisation probability, or the infection fatality ratio. We show how reproduction numbers and herd immunity levels depend on the underlying social dynamics.

## 1. Introduction

The fast spreading COVID-19 pandemic has destabilized the world during the past two years, forcing most countries into alternating periods of confinement and deconfinement. These sequential measures have attenuated the disease progression that otherwise would have increased exponentially, and thus prevented health care systems from getting destabilized [1, 2, 3, 4]. For example, the age-stratified hospitalization data for France during the past year exhibits how the national lockdowns at March 18, 2020 and October 30, 2020 have stopped the exponential pandemic growth (Fig. 1). With the availability of vaccines the pandemic seems under control if the population is progressively vaccinated. However, it remains the serious threat that current vaccines are not efficient to handle the constant emergence of new mutations, and the pandemic might get out of control again [5]. If no vaccines are available, for example at the beginning of a pandemic, monitoring and modeling tools are essential to predict and curtail the pandemic growth together with its consequences on the health care system [6].

**Figure 1:**
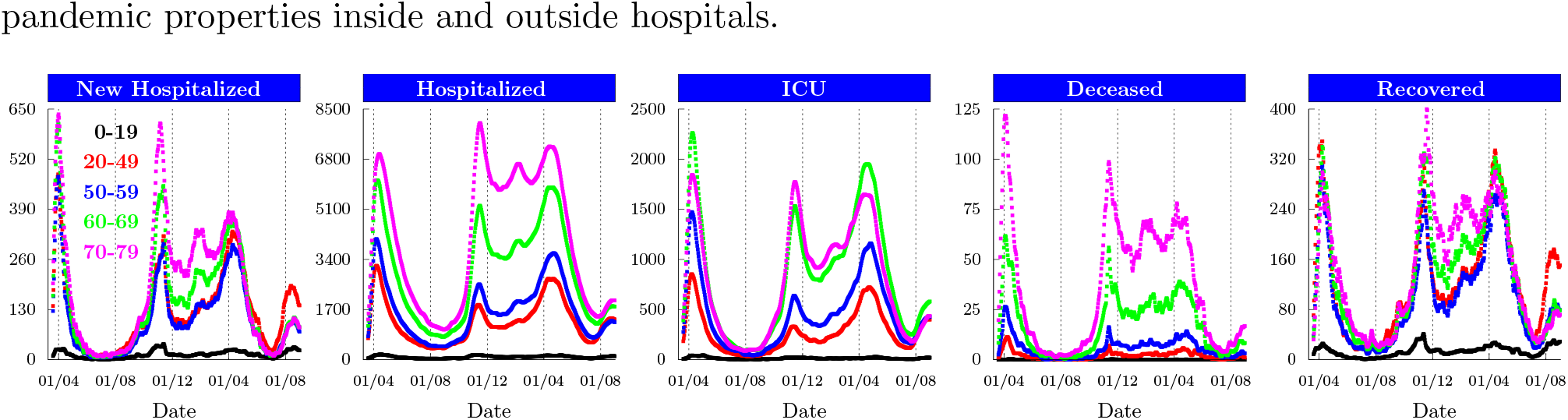
Age-stratified COVID-19 hospitalisation data for France. Daily number of patients newly hospitalised, hospitalised, in Intensive Care Units (ICU), deceased in hospitals and recovered patients that were released from hospitals. Data starting from March 18, 2020. Data sources are [35, 36].

During the early phase of the COVID-19 pandemic when vaccines were not available, the past experience has shown that surveillance, testing and severe social restrictions (non-pharmaceutical interventions, NPIs) attenuated the viral spread, which prevented hospitals from becoming overloaded. However, these restrictions not only came at a heavy economical cost [7], they also heavily disturbed social, school and academic life. Designing efficient social measures that not only curb the pandemic and exonerate the health care system, but also minimize social and economic impacts remains challenging [8]. In presence of a fast exponential pandemic growth characterized by a large reproduction number *R*_0_ ∼ 3, NPIs have to be taken as soon as possible because a small delay of only a few days can already significantly affect the hospital load [9].

In such circumstances, predictive models are indispensable to forecast the pandemic evolution and to timely anticipate the need of new social measures. During the past two years, a large variety of models have been developed to study the COVID-19 pandemic: continuous time SIR type compartmental models based on differential equations [10, 11, 12, 13, 14, 15, 16, 17]; SIR type models combined with Bayesian inference, SMC or MCMC methods [9, 18, 19, 20, 4, 21, 22, 23, 24, 25]; stochastic frameworks and agent based models [3, 26, 27, 2, 28, 1, 29, 30]; or deterministic discrete time compartmental models [31, 32]. The models were applied to study various aspects of the pandemic: to analyse the severity of the disease [19, 2], to estimate the impact of social measures [9, 1, 3, 26, 1, 33, 4], to study the early dynamics of the viral spread [20, 18, 31, 10], or to predict the effect of deconfinement measures [9, 3, 26, 18, 31, 34, 32].

Despite of these modelling efforts, it remains challenging to make reliable predictions during the early phase of a new pandemic due to incomplete knowledge, constant changes in social behaviour, or unexpected events. To cope with these challenges, we propose a data-driven dynamical framework that constantly adapts to new data in order to provide reliable near future predictions that can be used to control the pandemic evolution (Fig. 2). We calibrate the framework with a large variety of data (Fig. 2A): web accessible hospitalisation statistics (Fig. 1), clinical data from the Bichat hospital in Paris (Fig. 6), serology and positive testings (Fig. S2A), and surveys about social interaction matrices (Fig. 3D and Eq. 6 in the SI). We implemented an automated feedback and inference methods to constantly update model parameters to new data (Fig. 2A-B), and to subsequently readjust near future predictions of the pandemic and hospitalisation evolution (Fig. 2C-D). To perform dynamical simulations of the pandemic and hospital evolution, we devise a spatially coarse grained and time discrete compartmental model (Fig. 3). Besides the generic susceptible compartment, we have compartments that characterize the infection status of an infected person, for example asymptomatic, symptomatic, hospitalised or recovered (Fig. 3C). We stratify the population according to age (Fig. 3B), and we omitted persons older than 80 because many of them live in retirement homes where social interactions are different from the contact matrices used here. Beside the calendrical time, we introduce the compartmental time that counts the time since an infected person is in its current status, for example the time since an asymptomatic person became infected, or the time since a symptomatic person developed symptoms (Fig. 3A). We use transition probabilities that depend on the compartmental time to model the infection dynamics characterized by switchings between compartments. This is different from SIR type of models, where the dynamics is approximated with exponential transition rates. We use clinical data from a large Parisian hospital to compute hospital transition probabilities (Fig. 6). We implement fitting procedures to estimate changes in contact matrices due to NPIs from the data for the number of new hospitalisations (Fig. 4). We calibrate the model at national level in France until February 2021, since in this work we do not consider the impact of vaccinations and new viral strains. We use the causal dynamical simulations to study pandemic properties inside and outside hospitals.

**Figure 2:**
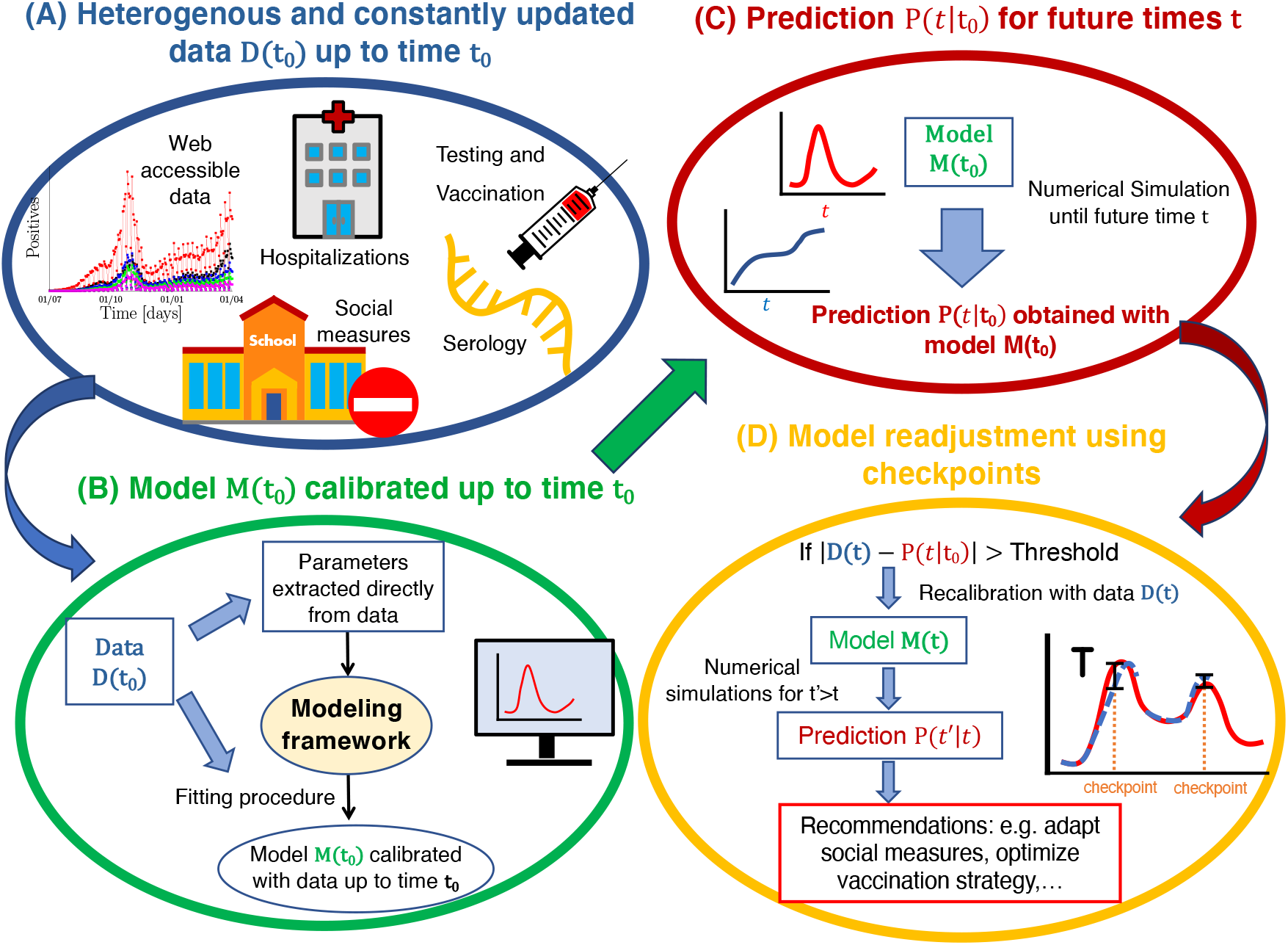
Automated data-driven checkpoint procedure to control a pandemic evolution. (A) A variety of constantly updated data is used to calibrate the model. (B) The model *M* (*t*_0_) is calibrated with data *D*(*t*_0_) up to time *t*_0_. (C) The calibrated model *M* (*t*_0_) is used to predict the pandemic and hospitalisation situation *P* (*t*|*t*_0_) at future times *t > t*_0_. (D) As time progresses, a checkpoint procedure constantly confronts the predictions *P* (*t*|*t*_0_) to the data *D*(*t*). If at some time *t* the discrepancy between model and data becomes larger than a threshold *T*, a recalibrated model *M* (*t*) is derived using the data *D*(*t*). The model *M* (*t*) is then used to generate new predictions *P* (*t*^*′*^|*t*) for times *t*^*!*^ *> t*. Based on these predictions, social measures and strategies are adjusted.

**Figure 3:**
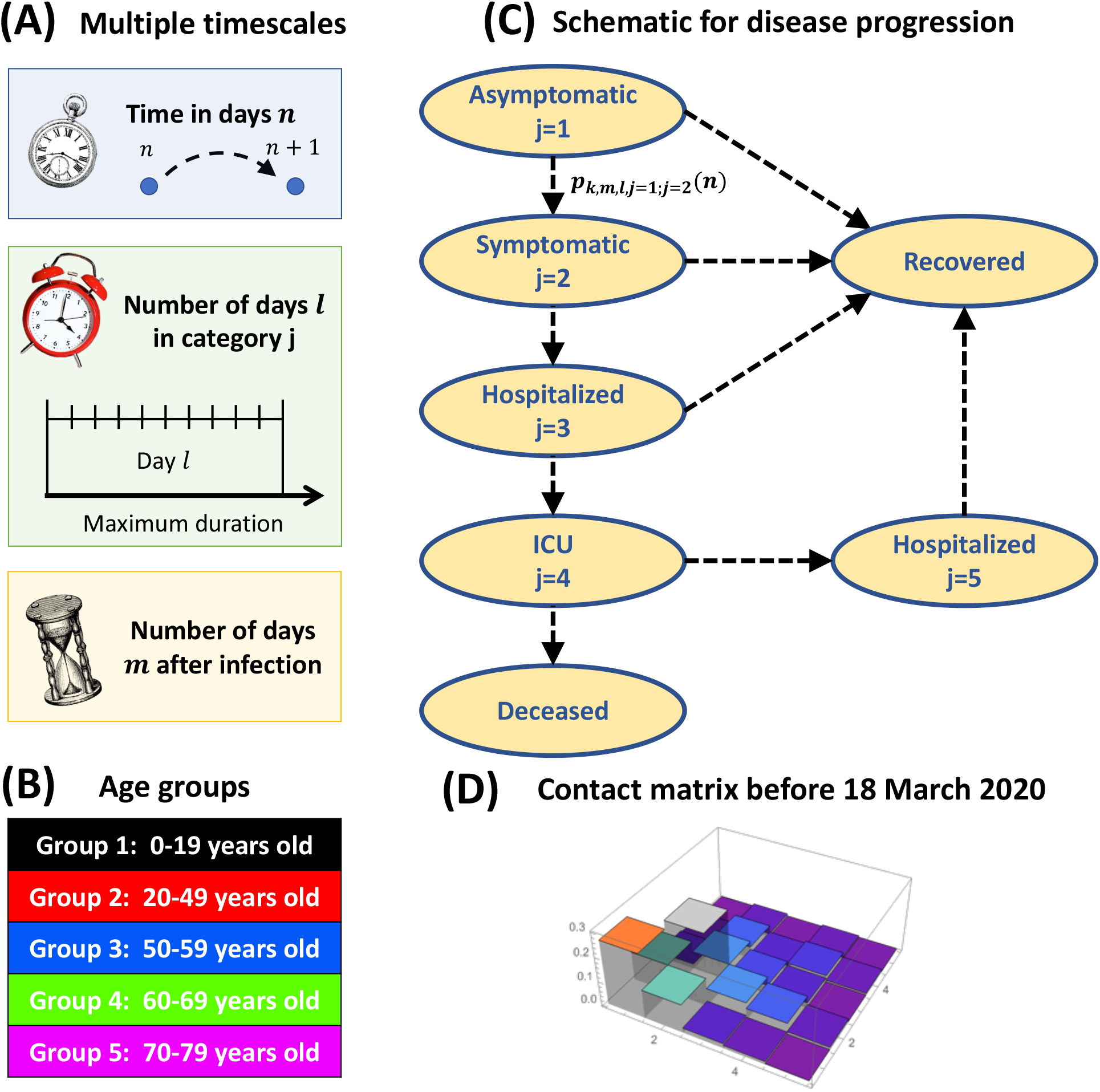
Time discrete and spatially coarse grained compartmental model. (A) Multiple time scales (in number of days): the calendrical time *n* since January 1, 2020, the compartmental time *l* since an infected joined its current compartment, or the time *m* since infection. (B) Definition of the five age groups considered in the model (labelled by the index *k*). We do not consider persons older than 80 because many of them live in retirement homes where the contact matrix is very different from the rest of the population. (C) Model for disease progression with transitions between 5 infection compartments that classify the infected population (labelled by the index *j*). We do now show the susceptible compartment. The transition probabilities 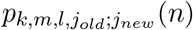 determine the switching dynamics from compartment *j*_*old*_ to *j*_*new*_. Infected persons only die in hospitals in Intensive Care Units (ICU). Recovered persons are differentiated depending on the compartment from which they recover. (D) Normalized contact matrix for France from [37] for the time period before the first lockdown at March 18, 2020.

**Figure 4:**
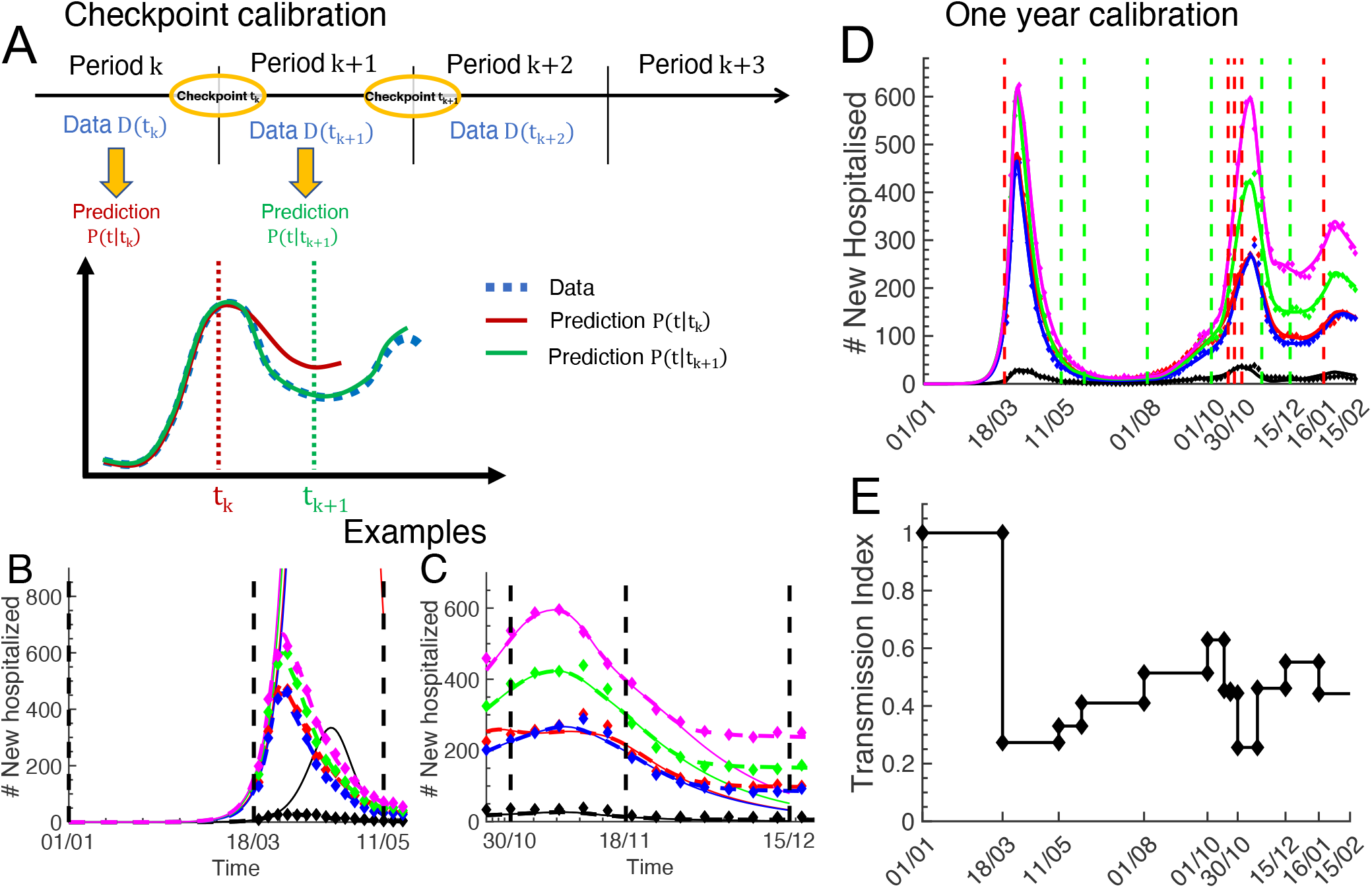
Sequential checkpoint calibration procedure with new hospitalisation data. The checkpoint times *t*_*k*_ correspond to social intervention times from Table 1. The new hospitalisation data (diamonds) in (B-D) is from Fig. 1. (A) Schematic of the sequential calibration procedure. The data for period k+1 ([*t*_*k*_, *t*_*k*+1_]) is used to fit the new contact matrix *c*_*a*;*a*_^*′*^ (*t*_*k*_) at time *t*_*k*_ that accounts for new social measures that are put into effect at *t*_*k*_. The model prediction *P* (*t*|*t*_*k*_) (red curve) computed with the unchanged contact matrix before deviates from the data for *t > t*_*k*_. The prediction *P* (*t*|*t*_*k*+1_) (green curve) is computed with the new contact matrix. (B-C) Two calibration examples. The predictions (solid lines) computed with contact matrices for the times before March 18 and November 18, 2020 deviate from the data (diamonds). The predictions computed with contact matrices that were adapted at March 18 and November 18 (dashed lines) reproduce the data. (D) Result of the sequential calibration procedure until February 2021. Model simulations (solid lines) are compared to the new hospitalisation data (diamonds). (E) The normalized transmission index 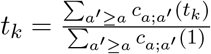 (see Material and Methods).

## 2 Materials and Methods

### 2.1 Web-available epidemiological and testing data, and hospital specific clinical data

To calibrate model parameters we use data from several sources, as described below.

**Table 1:** Checkpoint events where we changed the social contact matrix. Events that had a confining effect are marked with red color, deconfining events with green.

| Date | Event |
| --- | --- |
| 01/01/2020 | First day of simulation |
| 18/03/2020 | Beginning of the first lockdown |
| 11/05/2020 | Partial deconfinement and introduction of masks |
| 02/06/2020 | Further deconfinement |
| 01/08/2020 | Beginning of vacation period |
| 01/10/2020 | Beginning of the academic year |
| 17/10/2020 | Region specific curfew |
| 23/10/2020 | National curfew |
| 30/10/2020 | Beginning of the second lockdown |
| 18/11/2020 | Beginning of deconfinement |
| 15/12/2020 | Further deconfinement |
| 16/01/2021 | Installation of curfew at 18h |

#### Age-stratified hospitalisation data from the French governmental website

Age-stratified hospitalisation data is provided by the French Government (www.data.gouv.fr) since March 18, 2020. We used the databases *donnees-hospitalieres-covid19* and *donnees-hospitalieresclasse-age-covid19* to obtain daily number of patients hospitalised *H*(*n*), in intensive care (ICU), deceased in hospitals *D*(*n*), and recovered from hospitalisations *R*(*n*) (Fig. 1). To reduce fluctuations we smoothed the data with a Gaussian filter. We computed the daily number of new hospitalisations as *newH*(*n* + 1) = *H*(*n* + 1) − (*H*(*n*) − *R*(*n*) − *D*(*n*)).

#### Age-stratified testing data from the French governmental website

Age-stratified results from COVID-19 testings in laboratories and hospitals is provided by the French Government (www.data.gouv.fr) via the database *donnees-relatives-aux-resultats-des-testsvirologiques-covid-19*. The age stratified number of positive tested persons is shown in Fig. S2 in the SI. We also used published data about the prevalence of viral mutations (*donnees-de-laboratoirespour-le-depistage-indicateurs-sur-les-mutations*).

#### Age-stratified clinical data from the Bichat hospital

The Bichat hospital in Paris provided anonymized and age-stratified data collected between March 2020 and January 2021 that displays the hospitalisation history of 586 patients who all received ICU treatment. The data does not contain patients in the age group 0-19 years old. Statistics extracted from this data are shown in Fig. 6.

The clinical data comes from the OutcomeRea database that was declared to the *Commission Nationale de l’Informatique et des Libertés* (#999,262), in accordance with French law, and this study was approved by the institutional review board of Clermont Ferrand. Informed consent was not required because the study did not modify patients management and the data were anonymously collected.

### 2.2 Data based feedback algorithm to control the pandemic and hospitalisation evolution

We use a variety of data to update and calibrate model parameters (Fig. 2A): web accessible hospitalisation statistics (Fig. 1), clinical data revealing hospital procedures (Fig. 6), testing results (Fig. S2), and social contact matrices (Eq. 6 in the SI). We implement an automated feedback algorithm where predictions are compared to new data, new data is used to recalibrate parameters, and recalibrated parameters are used to generate improved predictions. The feedback procedure consists of three steps (Fig. 2B-D):

1. In the first step, the model *M* (*t*_0_) is calibrated with the data *D*(*t*_0_) up to time *t*_0_ (Fig. 2B). To adjust parameters we implemented fitting procedures. If new social measures are put into effect at time *t*_0_, the contact matrix is adjusted such that *P* (*t*|*t*_0_) takes into account the expected impact of these measures.
2. In the second step, the model *M* (*t*_0_) is used to forecast the pandemic and hospitalisation evolution *P* (*t*|*t*_0_) for times *t > t*_0_ (Fig. 2C).
3. As time progresses, a checkpoint procedure constantly confronts the prediction *P* (*t*|*t*_0_) to new data *D*(*t*) to verify whether the prediction is still conform with reality (Fig. 2D). If at time *t* the discrepancy between prediction and data becomes larger than a threshold *T*, |*D*(*t*)−*P* (*t*|*t*_0_) *> T*, a recalibrated model *M* (*t*) is derived with the data *D*(*t*) up to time *t*. With the model *M* (*t*) new predictions *P* (*t*^*′*^|*t*) are computed for times *t*^*′*^ *> t*. If the new predictions are no longer conform with the expectations, this might be an early indication that social measures have to be readjusted.

### 2.3 Multiscale modeling framework

The modeling framework that we developed implements the feedback algorithm specified above. The model is multiscale because it depends on calendrical and compartmental time, and on data acquired at national and single hospital level. Mathematical details are given in the SI. We developed a spatially homogeneous compartmental model with a discrete time-resolution of one day (Fig. 3A). We distinguish between 5 age groups (Fig. 3B) and 5 infection compartments (Fig. 3C). We use transition probabilities between these compartments to compute the disease progression (Fig. 3C). Social interactions are modelled with time-dependent contact matrices that account for national political interventions (Fig. 3D).

#### Multiple time scales to compute the disease progression

We distinguish between the calendrical time *n* that measures the number of days since January 1, 2020, and the compartmental time *l* that measures the number of days since an infected person joined his current infection compartment (Fig. 3A). The total time *m* since infection can be computed by adding the times spent in each compartment. For asymptomatic persons in compartment *j* = 1, the times *m* and *l* are identical. Transition probabilities might not only depend on the times *n* and *l*, but also on the time *m* since infection. However, since our hospital data only species the times *l* after hospitalisation (see Fig. 6), we use here only the times *n* and *l* for the modelling.

#### Population divided into five age-stratified groups

We classify the population into 5 age groups labelled by *k* = 1 … 5 (Fig. 3B): 0-19, 20-49, 50-59, 60-69 and 70-79 years old. The population in age group *k* is *N*_*k*_ = *N*_*tot*_*p*_*k*_, where *N*_*tot*_ = 67 millions and *p*_*k*_ = (24%, 36.3%, 13.1%, 11.9%, 10.2%) is from *https://www.statista.com/statistics/464032/distributionpopulation-age-group-france*. We do not consider persons older than 80 because many of them live in retirement homes where the interaction dynamics and contact matrix is very different from the rest of the population. For this fragile age group we suggest implementing a separate model.

#### Disease progression characterized by five infection compartments

Besides the susceptible population, to characterise the infected population we consider 5 infection compartments and transition probabilities that specify the disease progression (Fig. 3C). In the current implementation the transition probabilities depend only on the time *l*. A newly infected person starts in the asymptomatic compartment *j* = 1. From there it can either develop symptoms and become symptomatic (switching to *j* = 2), or remain asymptomatic and eventually recover. For a symptomatic person the disease either further deteriorates such that hospitalisation is needed (switching to *j* = 3), or it will recover without hospitalisation. A hospitalised patient in compartment *j* = 3 can be transferred to ICU (switching to *j* = 4), or it will eventually recover and be released from the hospital. In ICU, a patient can either die, or be transferred back to normal hospital (switching to *j* = 5). Finally, a patient in hospital compartment 5 will eventually be released and join the recovered population. In total, the model distinguishes between four types of recovered persons.

#### Infection dynamics leading to new hospitalisations with non-pharmaceutical interventions (NPIs)

Since the number of new infections is hidden, and the number of positive testings was not reliable enough throughout the beginning of the pandemic [27], we decided to use the data for the daily number of new hospitalisations to calibrate the infection dynamics leading to new hospitalisations (Fig. 1). We neglect the number of new infections that are possibly generated in hospitals. As a consequence, we can independently calibrate the model leading to new hospitalisations, and the model for the hospital dynamics. We use the model for new hospitalisations as input to calibrate the hospitalisation parameters of the model (see section 3.2).

In the following we discuss the procedures that we implemented to calibrate the model leading to new hospitalisations.

- As initial condition for the pandemic we fitted a seed of new infected persons in the age-group 20-49, since person from this age group are likely to have introduce the virus in France due to travailing. We fixed January 1, 2020 as initial date for simulations. We verified that changing this date does not much affect simulation results at later times because it can be compensated by changing the value of the initial seed.
- Initially, a new infected person is in the asymptomatic compartment *j* = 1. When the infection manifests itself with symptoms, the asymptomatic person switches to the symptomatic compartment *j* = 2. We assume that the now an alerted person takes precautions to avoid infecting others. Thus, in our implementation only infected persons belonging to the asymptomatic compartment spread the disease. We therefore consider a non-zero infectiousness only for the asymptomatic compartment (see Eq. 4 in the SI). However, we emphasize that the model is general and a non-zero infectiousness can be implement for other compartments if necessary, for example, to model new infections generated by symptomatic persons. We use an incubation period of 5 days [38], and we assume that a person is uniformly infectious within the time period *l* = 5 −11 days after the infection (Eq. 4 in the SI). We simplified and assumed that an infected person can develop symptoms with uniform probability within the period *l* = 6 − 11 days after the infection [39]. This gives two days (*l* = 5, 6) where an asymptomatic person transmits the disease before symptoms can appear (often referred to a pre-symptomatic transmission). Note that SIR models have to introduce a pre-symptomatic compartment to model an infected person that is asymptomatic but infectious. In contrast, with our time scale *l*, we can incorporate a time dependent infectiousness for the asymptomatic compartment without having to introduce a pre-symptomatic compartment.
- We fitted the age dependent probabilities to remain asymptomatic. To reduce the number of fitting parameters, we consider that the age groups 2-4 have the same probability to remain asymptomatic. For age group 1, we constrained the fitting range to values between 30% and 80%, for age groups 2-4 to 30% and 50%, and for age group 5 to 20% and 40%.
- Based on findings that the viral load is independent of age [40, 39, 41], we assumed that the infectiousness is independent of age. However, since it is unclear whether children and adolescents have the same infection dynamics as adults [42], we allowed that the susceptibility of group 1 to become infected is different from the other groups. We fitted the susceptibility of group 1 with the constraint that it is reduced by maximally 50% compared to the other groups (see Eq. 7 in the SI).
- We fitted the age-stratified probabilities for a symptomatic person to become hospitalised (Fig. S1). To constrain the fitting, we first fitted the data for new hospitalisations from the number of positive testings (Fig. S2) to extract age stratified probabilities that a positively tested person becomes hospitalised: we obtained 0.6%, 1.5%, 4.5%, 10.5%, and 22.8%. We used these values as lower limits for the probability that a symptomatic person becomes hospitalised, since there is also the possibility that persons have been tested positive multiple times, for example due to travelling, or that a person become hospitalised without being tested before. Due to reduced travelling and social interactions, we assumed that a person in the age group 70-79 is tested only when showing symptoms. We therefore reduced the fitting parameters by using the value of 22.8% for group 5 as a fixed anchor (Fig. S1A in the SI). We further reduced fitting parameters by assuming that the distribution, which specifies at which day *l* a persons becomes hospitalised after showing symptoms, is age independent (Fig. S1B in the SI).
- We modified contact matrices to account for multiple governmental interventions that affected social interactions. Table 1 specifies the dates when new social restrictions have been put into effect (red entries), or restrictions have been relieved (green entries). We added two additional dates beginning of August and October to account for the beginnings of summer vacation and new academic year. The dates in Table 1 correspond to checkpoint times *t*_*k*_ where contact matrices were modified (all other model parameters were kept unchanged). We divided the past year into consecutive time periods that are delimited by these checkpoint times. For example, period_1_ ([*t*_0_, *t*_1_]) corresponds to the time from January 1, 2020 until March 18, 2020 when the first national lockdown was imposed, and period_2_ ([*t*_1_, *t*_2_]) is the lockdown period until May 11, 2020. At each time *t*_*k*_ we fitted a new contact matrix *c*_*a*;*a*_^*′*^ (*t*_*k*_) (15 fitting parameters) using the new hospitalisation data for period_*k*+1_ ([*t*_*k*_, *t*_*k*+1_]) (Fig. 3A).

We extracted the contact matrix during period 1 before the first lockdown in March 2020 from [37] (see Eq. 6 in the SI). We fitted the contact matrix for the lockdown period (period 2). To constrain the lockdown contact matrix parameters (15 parameters), we made the following assumptions about the impact of the lockdown measures (see also [43, 3]): contacts between group 1 and groups 4 and 5 are reduced by at least 90%, since grandparents could no longer be visited; due to school closure, the intra-group contact of group 1 is reduced by at least 70%; all the remaining contact parameters are reduced by at least 40%.

Since new hospitalisation data was not available for period 1, we simulated period 1 and 2, and we used the new hospitalisation data from period 2 for the fitting. We fitted the unknown infection parameters (initial seed, fraction asymptomatic, infectivity, susceptibility of age group 1, probabilities for symptomatic to become hospitalised; see also the paragraph further down where we summarize the fitted values for the infection parameters) and the lockdown contact matrix. Our inference methods are such that any model parameter can be fitted. To reduce the number of consecutive fitting parameters, we adopted a recursive fitting procedure: We first generated a lockdown contact matrix that we used as input, and we fitted the infection parameters by minimizing the mean squared error between simulation and new hospitalisation data for period 2. Then we used the fitted infection parameters as input, and in a second round we fitted the lockdown contact matrix with the new hospitalisation data for period 2. Then we used the fitted lockdown contact matrix as input and we refitted the infection parameters, etc. We recursively performed this procedure until the change in the mean squared error became lower than a chosen threshold value.

For the time after the lockdown (after May 11, 2020), we fitted only contact matrices and we kept the infection parameters unchanged. Since masks were largely unavailable before May 11, the pandemic declined during the first lockdown as a consequence of a reduction in social contacts. In contrast, with the availability of masks, the infection dynamics further depends on a reduced infectivity of social interactions due to mask wearing [44]. However, because a reduction in the infectivity parameter *β*(*n*) due to masks has the same effect as a uniform reduction of the contacts matrix (Eq. 10 in the SI), we adopted the following fitting procedure after May 11: 1) we kept *β*(*n*) constant and fitted only contact matrix parameters; 2) if the social measures had a deconfining character, we used the preceding contact matrix as lower limit, otherwise as upper limit. Consequently, the fitted contact matrices after from May 11, have to be considered as effective matrices that also comprise the effect of mask wearing. With this method we sequentially calibrated the model to the new hospitalisation data until February 2021 (Fig. 4D; fitting results are solid lines, diamonds are data; vertical red dashed lines indicate confining events, green dashed lines deconfining events). To visualise these changes, we defined the transmission index 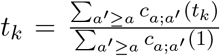, and we show how this value evolved due to mask wearing and social measures (Fig. 4E). The index is normalized to one for the initial uncontrolled phase of the pandemic. The strong reduction of the transmission index by almost 80% during the first lockdown after March 18, 2020 is entirely due to contact reductions, whereas the values at later times comprise the combined effect of mask wearing and contact reduction.

Without changing the contact matrix at March 18, 2020 the pandemic would have continued to grow exponentially (Fig. 3B, solid curves). In contrast, with the lockdown contact matrix the model reproduces the pandemic decline during the lockdown (Fig. 3B, dashed curves). As another example that shows the impact of contact matrix changing, we consider October 30 where a second national lockdown has been imposed that lasted officially until November 28. However, we could not satisfactorily reproduce the new hospitalisation data by assuming the same contact matrix between October 30 and November 28 (Fig. 3C, solid curves). Instead, a much better fit was obtained by adapting the contact matrix already around 10 days before the official end of the lockdown (Fig. 3C, dashed curves), suggesting that the social interaction dynamics started to change already before the official end of the lockdown.

#### Summary of fitted infection parameters

The initial seed of infected in age group 2 at January 1, 2020 is 11. The fraction of asymptomatic is 80%, 46.15%, 46.15%, 46.15%, and 20%. The values 80% and 20% correspond to upper and lower limits. The susceptibility of age group 1 was reduced by 37% compared to the other groups. The value for the infectivity *β*(*n*) was 2.63 ×10^7^ (Eq. 7 in the SI). This value determines the exponential growth during the initial phase, and depends on the normalisation of the contact matrix. The probability that a symptomatic person becomes hospitalised is given in Fig. S1 in the SI.

## 3 Results

Although there is regional heterogeneity of the infection spread throughout France [32, 3, 45], we adopted a coarse grained approach to model the pandemic evolution at national level. We split the model into two parts: In the first part we model the infection dynamics leading to new hospitalisations, and in the second part we use the new hospitalisations to predict the hospital workload.

### 3.1 Infection dynamics with multiple social interventions

Multiple governmental interventions during the past year affected social interactions, which we accounted for by adapting the contact matrix of the model (see Materials and Methods). The latest contact matrix accounts for the social measures from January 16, 2021. We did not derive contact matrices for the time after January 20201 because the model does not include the effect of vaccinations and viral mutations. We simulated the pandemic evolution during 2021 to show the consequences of a scenario with persisting social restrictions from January 2021 (Fig. 5; the vertical dashed lines indicate that the model has been calibrated to data until February 15, 2021).

**Figure 5:**
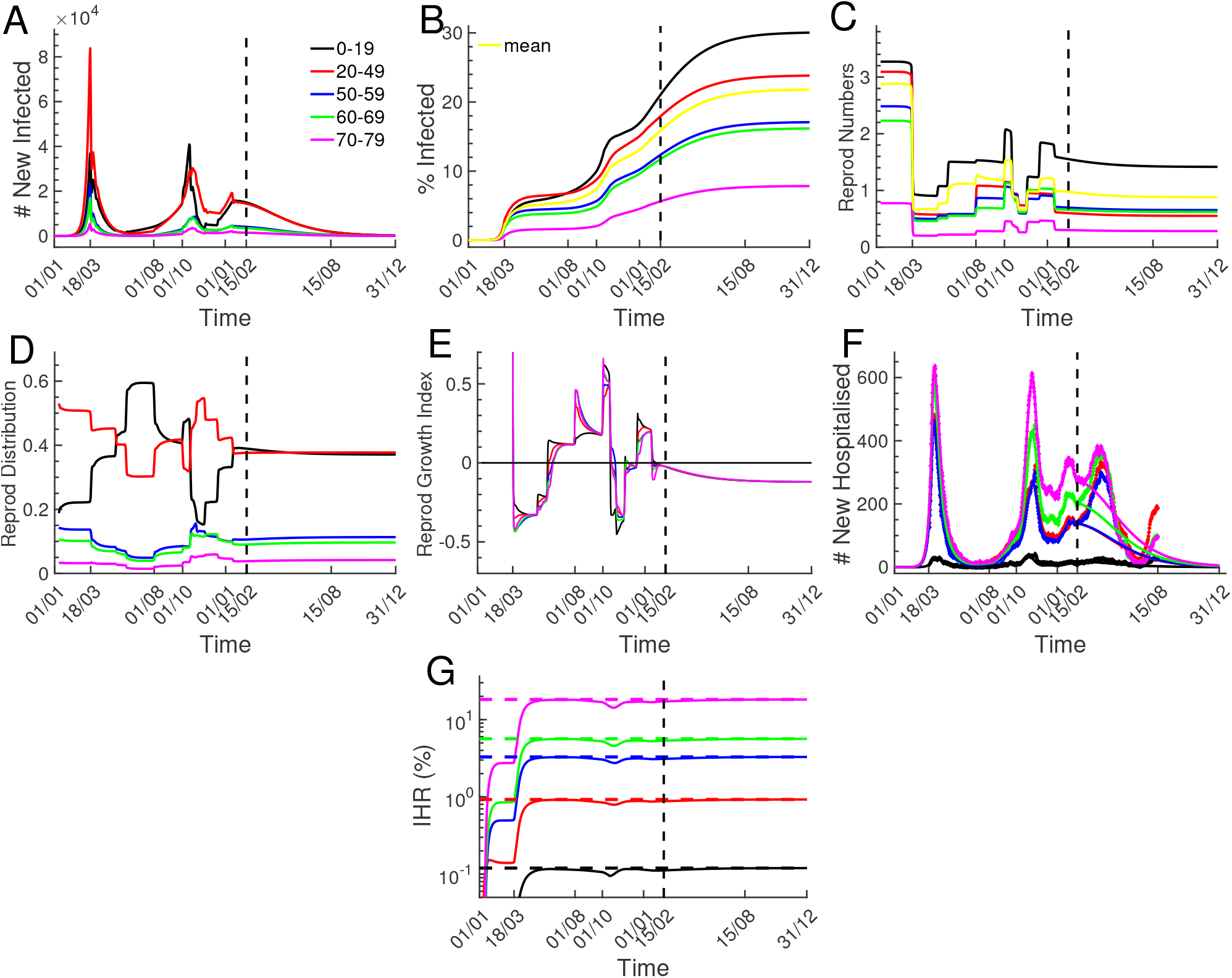
Infection dynamics with multiple social interventions. Simulations from January 1, 2020 until December 31, 2021 performed with the model that has been calibrated to data until February 15, 2021, as indicated by the vertical dashed lines. Because the model does not account for new viral strains and vaccinations, the simulations after February 2021 show the pandemic evolution for a hypothetical scenario without new viral strains and vaccinations, and social restrictions from January 2021 that persist throughout the year. (A) New infections per age group. (B) Fractions of infected population per age group, and mean fraction of infected population (yellow curve). (C) Age group specific effective reproduction numbers *R*_*k*_(*n*) (Eq. 12 in the SI). The value *R*_*k*_(*n*) gives the average number of new infections that will be generated by a person that becomes newly infected at day *n* in group *k*. The yellow curve represents the mean reproduction number *R*(*n*) (Eq. 13 in the SI). (D) Reproduction distributions showing in which age group new infections will be generated (Eq. 14 in the SI). (E) Reproduction growth indices (Eq. 15 in the SI). A positive value indicates that the number of new infections will increase in the specific age group. (F) New hospitalisation simulations (solid lines) are compared to the data from Fig. 1. (G) Probability that an infected person becomes hospitalized (Infection Hospitality Ratio, IHR). The IHR values are obtained by dividing the cumulated new hospitalisations from (F) with the cumulated new infections from (A). The values 0.12%, 0.92%, 3.3%, 5.6% and 18.2% marked by the dashed lines are the probability for an asymptomatic to become symptomatic multiplied by the probability for a symptomatic to become hospitalised.

The simulated number of new infected per age group exhibit three prominent peaks in March 2020, October 2020 and January 2021 when the pandemic was not under control (Fig. 5A). Interestingly, the various age groups contribute very differently to new infections: the dominant contributions come from age groups 0-19 and 20-49 due to their large number of interactions (contact matrix, Eq. 6 in the SI), whereas infected persons in the age group 60-79 contribute only little to spread the disease (Fig. 5A). The simulation reveals that with the social restrictions from January 2021 the pandemic would have declined during 2021 if no viral mutations had appeared.

From the number of new infections, we computed the age dependent fraction of infected population (Fig. 5B). At May 11, 2020 around 5% of the population had been infected, which increased to around 13.6% until January 16, 2021 (Fig. 5B, yellow curve). However, whereas by January 16, 2021 only around 5% of the population in the age group 70-79 has been infected (Fig. 5B, magenta), this value increases to around 18% for the age group 0-19 (black).

To characterize the pandemic growth, we computed age-stratified effective reproduction numbers *R*_*k*_(*n*) that represent the mean number of new infections that will be generated by a person that becomes newly infected at day *n* in age group *k* (Eq. 12 in the SI). By averaging over age groups we further defined the mean reproduction number *R*(*n*) (Eq. 13 in the SI). We found that the reproduction numbers *R*_*k*_(*n*) are strongly age and time dependent (Fig. 5C). This reflects the impact of multiple social measures and the age stratification of interaction frequencies. For example, the reproduction number for age group 70-79 is always smaller than one due to the low interaction frequency, whereas it is almost always above one for age group 0-19 (Fig. 4C, magenta vs black curve).

To characterise how new infections are distributed among age groups, we defined the reproduction distribution *RD*_*k*_(*n*) that specifies the fraction of new infections generated in age group *k* (Eq. 14 in the SI). We found that more than 80% of new infections are generated within group 1 and 2 (Fig. 4D).

Because the reproduction numbers *R*_*k*_(*n*) provide only information about the number of infections that will be generated by an infected person from group *k*, they do not reveal whether the new infections in group *k* will decline or increase. To characterise these changes, we introduced reproduction growth indices *RGI*_*k*_(*n*) (Eq. 15 in the SI) that measure the change in new infections per age group (Fig. 4E). A positive values indicates that the pandemic grows. Interestingly, although the reproduction number for age group 0-19 is larger than one during 2021, the growth index for this group is negative. This shows that infected persons in this age group spread the disease to other groups.

In Fig. 5G we compare the new hospitalisation simulation (solid lines) to data from Fig. 1 (diamonds). Up to February 15, 2021, Fig. 5G is identical to Fig. 4D. The new hospitalisation data has a forth peak beginning of April 20201, which is probably due to the effect of more infectious viral strains. The sharp decline in April and May is probably a combined effect of vaccinations and social restrictions from March 20201. In contrast, with the contact matrix from January 2021 and without new viral strains and vaccinations, the simulation predicts that the number of new hospitalisations would have gradually declined during 2021.

Finally, we computed the Infection Hospitality Ratio (IHR) (by dividing the cumulative number of new hospitalisations with the cumulative number of new infections from (A) Fig. 5G, continuous lines). We further computed analytic IHR values (Fig. 5G, dashed lines) by multiplying the probability for infected persons to become symptomatic with the probability for symptomatic to become hospitalised (the probabilities are specified in the Materials and Methods), and found the age group dependent values 0.12%, 0.92%, 3.3%, 5.6%, and 18.2%. Thus, an infected person in age group 70-79 is around 150 times more likely to become hospitalised compared to a person in age group 0-19. Interestingly, the transient plateau values in Fig. 5G before March 18,2020 differ from the analytic predictions during the exponential growth phase because of the time delay between new infection and new hospitalisation.

### 3.2 Hospitalisation dynamics

Predicting the hospital workload from the number of new hospitalisations requires statistics at single hospital level. Because such data is not provided by the French governmental websites, we used age stratified clinical data from the large Bichat hospital in Paris to extract hospital transition probabilities (Fig. 3). With the clinical data, we derived age stratified distributions specifying the day *l* at which patients are transferred to ICU, leave ICU, die in ICU, and are released from hospital (Fig. 6A-D, dashed lines). We further extracted the probability that a patient dies in ICU (Fig. 6E). Due to the small sample size per age group, the distributions show large fluctuations (Fig. 6A-D, dashed lines). We fitted the data with the discrete Gamma distribution (Eq. 26 in the SI) to smooth the curves, which reveal more clearly difference between age groups (Fig. 6A-D, solid lines). However, we found that using the smooth or fluctuating distributions had only a minor impact on predicting the hospital workload (see next paragraph). Finally, we computed the average number of days patients spend in compartments *j* = 3, 4, 5 before making transition to a new compartment (Fig. 6F).

**Figure 6:**
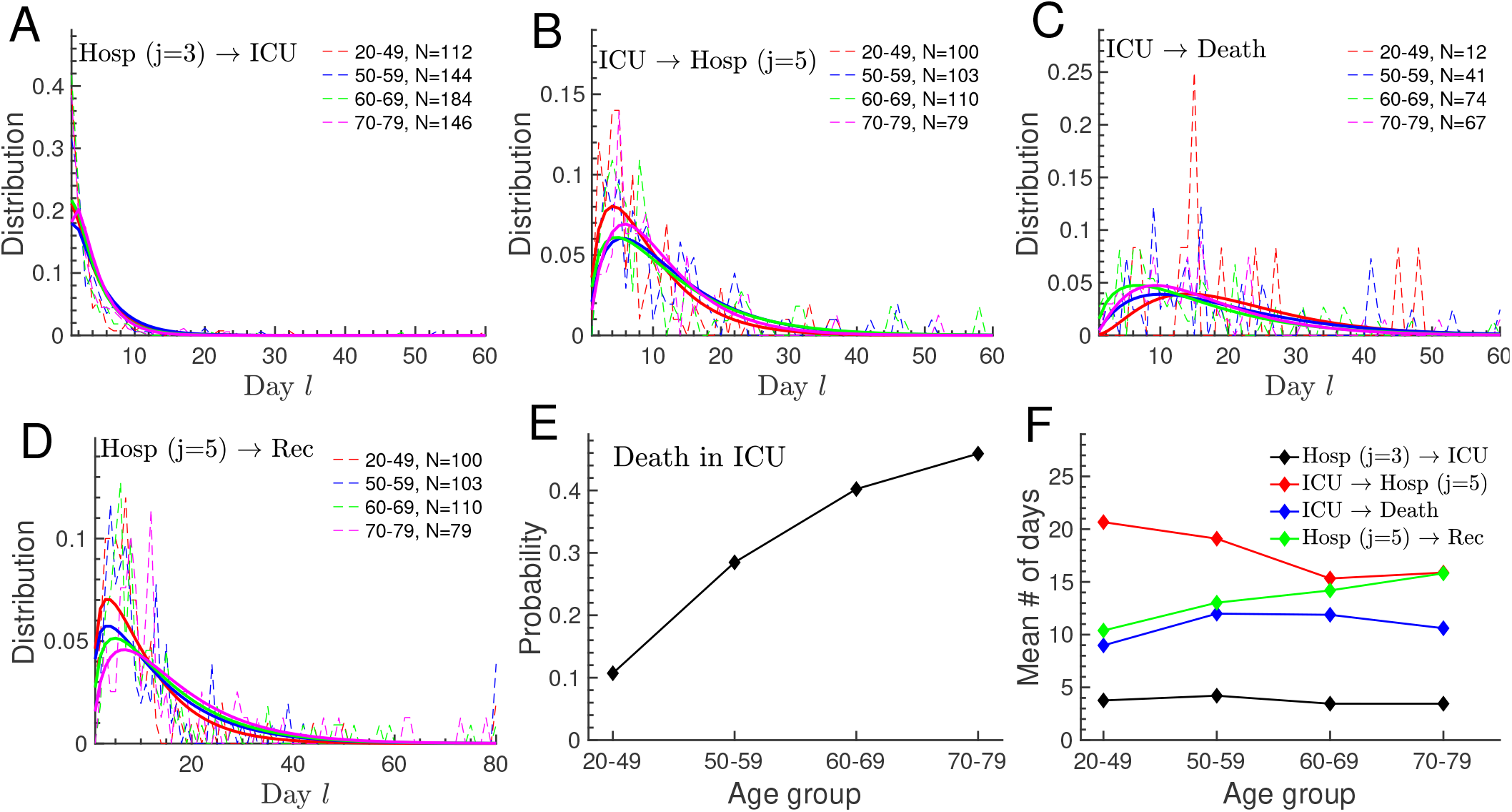
Hospital statistics and transition probabilities from clinical data. Age stratified statistics were computed with clinical data of 586 patients from the Bichat hospital in Paris (see Materials and Methods). All patients received ICU treatment and all were older than 20 years. (A-D) Switching distributions as a function of the number of days *l* after joining the compartment. We truncated *l* to a sufficiently large value *l*_*max*_ = 60 for *j* = 3, 4, and *l*_*max*_ = 80 for *j* = 5. Fluctuating dashed lines show the data, and smooth solid lines are fits obtained with the discrete Gamma distribution from Eq. 26 in the SI. The parameter *N* is the number of patients that contributed to the distribution. The individual panels display the distributions to be transferred from hospital (*j* = 3) to ICU (*j* = 4) (A), from ICU to hospital (*j* = 5) (B), to die in ICU (C), and to be released from hospital (D). (E) Probability to die in ICU. (F) Mean number of days before switching computed with the dashed distributions from (A-D).

In the subsequent step, we applied the transition probabilities from Fig. 6 to simulate the hospital workload in France based on the evolution of new hospitalisations (Fig. 5F). This implicitly assumes that procedures in the Bichat hospital are not too different from the national average. Unfortunately we could not extract the probability to be transferred to ICU, the distribution when patients without ICU treatment are released from hospital, and there was no data for age group 0-19. To compensate for this missing data, we proceeded as follows: we used for age group 0-19 the distributions for age group 20-49 shown in Fig. 6A-D, since we expect similarities between these two age groups, and since the exact shape of the distribution has only a minor impact compared to the overall switching probability (note that distributions are normalised to one); the remaining unknown probabilities were estimated by fitting for each age group the hospitalisation data for France until February 2021 (Fig. 1). We concurrently fitted the evolution of patients hospitalised, in ICU, that died in ICU, and released from hospital. For example, for the age group dependent probabilities to be transferred to ICU, we obtained the values 15.2%, 19.5%, 26.0%, 35.7%, and 31.4%.

Finally, we simulated the hospital workload from January 2020 until December 2021, which we compared to data from Fig. 1 (Fig 7A-D, simulations (solid lines) versus data (diamonds)). The simulated new hospitalisation from Fig. 5F were used as input. With hospital parameters from the Bichat data we could satisfactorily reproduce most of the hospitalisation dynamics in France until February 2021. The simulated number of deceased persons in the age group 70-79 is significantly reduced compared to the France data (Fig 7C), which might indicate that the Bichat hospital has lower probability to ie in ICU compared to the national average. We could have reduced this discrepancy by fitting the probability to die in ICU for this age group. We obtain larger discrepancies between simulation and data for the number of hospitalised patients (Fig 7A) due to differences in the daily number of recovered persons that accumulate over time (Fig 7D). The simulation further suggest that the social contact restrictions from January 2021 would have been sufficient to end the pandemic had no new viral strains emerged. Lastly, we computed the age dependent Infection Fatality Ratio (IFR) by dividing the cumulated numbers of deceased persons and new infections (Fig 7E, solid lines). The analytical values 0.0005%, 0.019%, 0.24%, 0.81%, and 2.62% are obtained by multiplying the probabilities to become symptomatic with the ones for symptomatic to become hospitalised, for patients to be transferred to ICU and to die in ICU (Fig 7F, horizontal dashed lines). For the mean IFR ratio we find a value of 0.2%. Remarkably, the probability to die after infection in the age group 70-79 is more than 5000 times higher compared to the age group 0-19.

**Figure 7:**
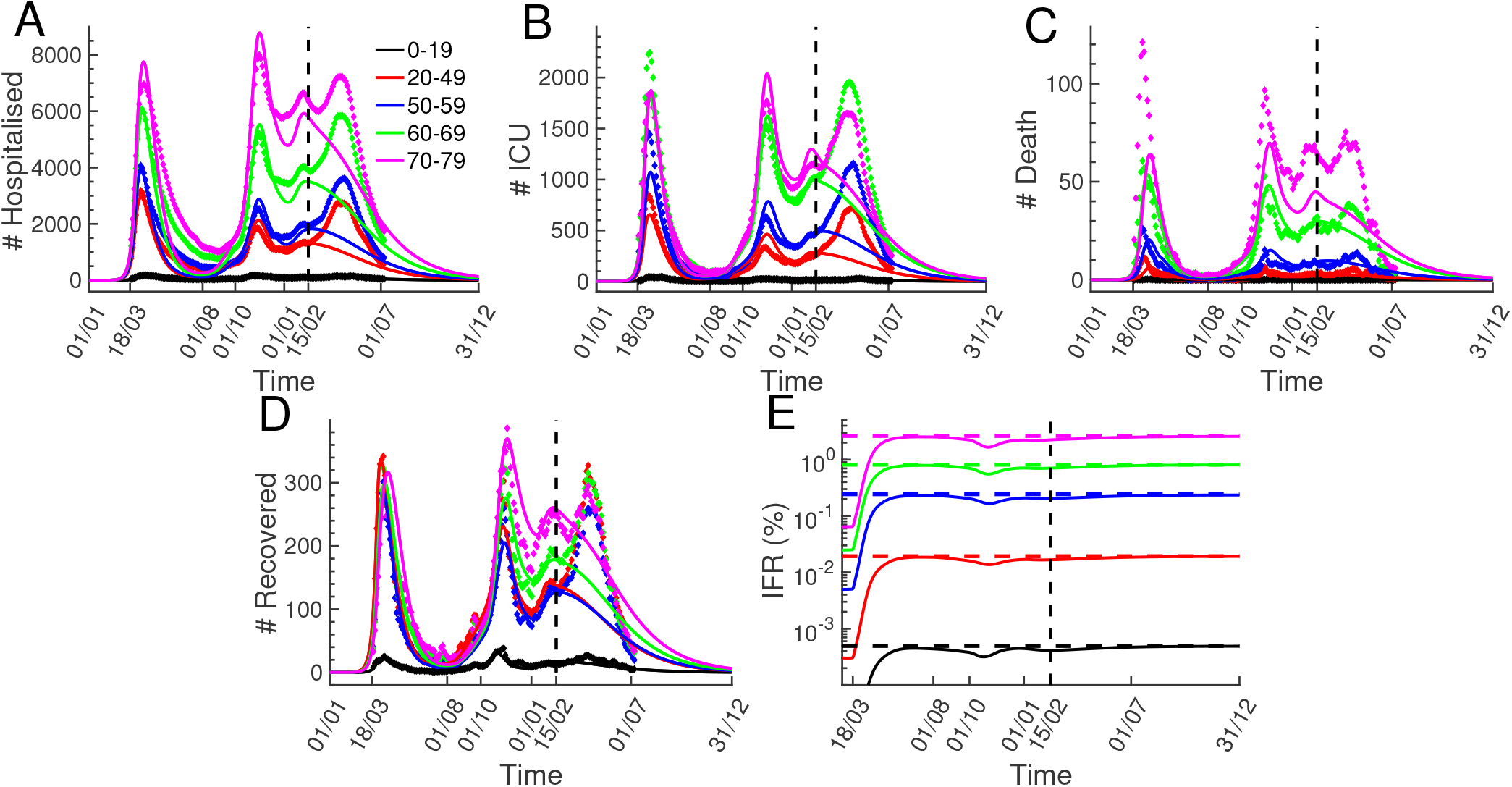
Hospitalisation dynamics. Simulations of the hospitalisation evolution (solid lines) are compared to data from Fig. 1 (diamonds). Simulations were performed with the new hospitalisation simulations from Fig. 5F as input, and with hospital parameters extracted from the clinical data in Fig. 6). The simulations after February 2021 show the hospitalisation progression in a scenario where the social restrictions from January 2021 would have persisted throughout 2021 without the impact of new viral strains and vaccinations. (A-D) Evolution of patients hospitalised (A), in ICU (B), deceased in ICU (C), and recovered patients that were released from hospitals (D). The simulated recovered are the sum of patients released from hospital compartments *j* = 3 and *j* = 5. (E) Age group dependent Infection Fatality Ratio (IFR) computed by dividing the cumulated deceased from (C) with the cumulated new infections from Fig. 5A. The dashed lines indicate the analytic values 0.0005%, 0.019%, 0.24%, 0.81%, and 2.62% computed with the probabilities for asymptomatic to become symptomatic, for symptomatic to become hospitalised, to be transferred to ICU, and to die in ICU. For the mean IFR we find 0.2%.

## 4 Discussion

During the early phase of a pandemic when vaccines are not available, social restrictions are key to prevent an exponential growth and a possible collapse of the health care system. In such conditions, realistic models based on a large variety of data are needed to timely test and optimize NPIs in a constantly changing pandemic environment. In this work, we developed a data-driven multiscale modelling framework together with methods to continuously adapt parameters to new data in order to make reliable near future predictions (Fig. 2). Key features of the framework: 1) age-stratified compartmental model with two time scales, calendrical and compartmental time (Fig. 3); 2) nonexponential transition probabilities that depend on the compartmental time (Fig. 3); 3) use of clinical data to extract hospital transition probabilities (Fig. 6); 4) development of fitting methods to infer model parameters and to adjust contact matrices to NPIs (Methods and SI); 5) regular checkpoint procedures that accommodate the model to new data and adjust near future predictions (Fig. 4).

To show the relevance of our work, we calibrated the framework to study the COVID-19 pandemic at national level in France with multiple social interventions until February 2021. We did not consider data beyond this time because the model does not include vaccinations and multiple viral strains, which changed the pandemic evolution afterwards. We implemented 5 age groups, 5 infection compartments, one compartment for deceased and 4 for recovered persons (Fig. 3B,C). Besides the calendrical time *n*, we consider the number of days *l* since an infected person joined its current compartment (Fig. 3A). Since transition probabilities are functions of the compartmental time *l*, we do not implement compartments like exposed, paucisymptomatic or prodromic, which are necessary in SIR type of models based on exponential switching rates. For example, instead of having an exposed compartment, we simply set the infectivity parameters to zero for the time *l <* 5 after infection (Eq. 4 in the SI). We classified non-hospitalised infected people into asymptomatic and symptomatic, similar to the dual classification from [46]. The difference between asymptomatic and symptomatic persons is that the latter are aware of their infection and take precautions to avoid infecting others. Infected persons with unnoticed symptoms therefore belong to the asymptomatic compartment. Consequently, we consider that only persons belonging to the asymptomatic compartment spread the disease, in agreement with the prevalent role of undocumented and silent transmissions [47, 48, 28, 23].

We developed constrained fitting procedures to infer model any unknown model parameter from data. We calibrated the infection parameters that are needed to simulate new hospitalisations by fitting data for new hospitalisations (Fig. 4D) and data for positive testings (Fig. S1). We sequentially fitted changes in contact matrices as a function of the underlying NPIs until February 2021 by minimizing the mean squared error between new hospitalisations data and simulation (Fig. 4D). For the age groups 20-69 and 70-79 we fitted that 46% and 20% of the infections remain asymptomatic, respectively, values that are within the large published range of 15% to 60% [49, 50, 39, 3, 26, 39, 31]. In contrast, for the age group 0-19 we fitted a surprisingly high value around 80% due to the following reasons: 1) the age group 0-19 is involved in around 42% of all contacts, comparable to 57% for age group 20-49 (Eq. 6 in the SI); 2) the probability for a symptomatic person in the age group 0-19 to become hospitalised is around 0.6%, which is reduced only by a factor 3 compared to the age group 20-49 (Fig. S1); 3) new hospitalisations in the age group 0-19 are reduced by a large factor 16 compared to 20-49 (Fig. 1). As a consequence, to obtain such a drastic reduction in the number of new hospitalisations, the probability to remain asymptomatic has to be much higher in the age group 0-19 compared to 20-49. A much higher asymptomatic fraction for children and adolescents is in agreement with more asymptomatic and mild infections, and fewer symptoms [51, 52, 53, 54, 55]. We further fitted a 37% reduced susceptibility to infections in age group 0-19, compatible with a reduced susceptibility for children and adolescents [56, 57, 52]. We tested scenarios where the susceptibility to infections is independent of age, and found that the fraction of asymptomatic in the age group 0-19 needs to be even higher around 90% (not shown). In summary, these results indicate profound differences in the disease progression between children and adults.

The simulations revealed that by May 11, 2020 around 5% of the French population had been infected, a value that increased to around 13.6% by January 16, 2021 (Fig. 5B, yellow curve). These values are consistent with 5.7% and 14.9% obtained with a completely different method based on serological data and a deconvolution procedure [45]. We estimated that the fraction of infected population is strongly age dependent: by January 16, 2021 only 5% of the population has been infected in the age group 70-79, contrary to 17% in the age group 0-19, similar to the values from [45]. We find that effective reproduction numbers are strongly age dependent (Fig. 5C), which is often ignored [8]. Since reproduction numbers depend on social interactions, there is no absolute value for the level of herd immunity [32, 12]. For example, with the contact matrix from January 2021 the herd immunity level would be around 21% (Fig. 5B, yellow curve). The effective reproduction numbers *R*_*k*_ count the number of infections that a person in age group *k* will generate (Eq. 12 in the SI), but they do not specify how these infections are distributed among age groups, and whether the infection level in a specific group will decline or increase. To quantify these properties, we introduced a reproduction distribution *RD*_*k*_ (Eq. 14 in the SI) and a reproduction growth index *RGI*_*k*_ (Eq. 15 in the SI). By computing *RD*_*k*_ we found that more than 80% of new infections are generated within age groups 0-19 and 20-49 (Fig. 4D). The *RGI*_*k*_ values revealed that the infection level can decline in an age group (Fig. 5E, black curve) although the reproduction number is above one (Fig. 5C, black curve). We computed infection hospitality ratios (IHR) and found the age group dependent values 0.12%, 0.92%, 3.3%, 5.6%, and 18.2% (Fig. 5G), comparable to estimations from [19, 45]. These values reveal that a person in age group 70-79 is around 150 times more likely to become hospitalised compared to person in age group 0-19.

We simulated the hospital workload with the simulations for new hospitalisations as input (Fig. 5E). Since the French governmental websites do not provide data at single hospital level to quantify hospital procedures, we used clinical data from the Bichat hospital in Paris to estimate hospital transition probabilities (Fig. 6). Assuming that the Bichat hospital can be used as a template for France, we used these parameters at national level. Because the Bichat data was incomplete to extract all hospital parameters, we quantified the remaining ones by fitting the national data until February 2021 (Fig. 7A-D). With this mixed procedure we could satisfactorily reproduce the hospitalisation dynamics (Fig. 7). We further tested a different method where we did not use the Bichat data, but we fitted all hospital parameters. As expected, this yielded a better agreement with the national data and slightly different parameter values. For example, the fitting procedure yielded a slightly higher probability to die in ICU for age group 70-79, which would have reduced the discrepancy between data and simulation for this age group in Fig 7C. Nevertheless, since we think that the use of clinical data is the appropriate way to estimate hospital parameters, we focused on the results obtained with this method. Assuming that infection parameters would not have changed after February 2021, we extended the simulation until the end 2021. This revealed that the social contact restrictions from January 2021 would have been sufficient to end the pandemic if no new viral strains had emerged. By combining the infection and hospitalisation parameters, we computed age group specific IFR values of 0.0005%, 0.019%, 0.24%, 0.81%, and 2.67%, with a mean 0.2%, consistent with results from [19, 47, 58, 59, 50, 22]. The COVID-19 mortality is strongly age dependent, for example, an infected person in the age group 70-79 has a more than 5000 times higher decease probability compared to a person in the age group 0-19.

There are several possibilities to extend and improve this framework in future work. To comply with the actual pandemic situation, it will be necessary to extend the model to handle vaccinations and multiple viral strains. It is conceptually straightforward to include vaccinated persons by further stratifying the population besides the age criteria. To handle multiple viral strains that have different infection and hospitalisation properties, separate models for each strain have to be calibrated and then coupled to make predictions. To better account for spatial inhomogeneities, separate models at national, regional or single hospital level could be implemented, or a spatially resolved social dynamics could be considered [60, 25]. Mobility and contact network data could be used to further constrain the fitting of contact matrices, and to acquire a more precise understanding of how NPIS correlate with changes in contact matrices [30]. For example, to design NPIs that control a future pandemic, one could generate artificial new hospitalisation fluxes that do not saturate hospitals, then use the fitting methods to estimate the contact matrices that generate such fluxes, and finally identify the NPIs that result in such contact matrices. Since in this work we focused on the calibration procedure, and we used a cost function based on the mean squeed error between data and simulations for the fittings. However, by choosing a different cost function one can design NPIs that achieve specific aims, for example, minimizing the number of new infections for elderly persons, or reducing economic cost. The goal will be to develop a comprehensive framework and software to control and manipulate the current pandemic, but also to provide a ready-to-use template that can be quickly activated in case of a new pandemic when vaccinations are not yet available.

## Data Availability

Data is available upon request to the authors.

## Supplementary Information

### A Supplementary Figures

**Figure S1:**
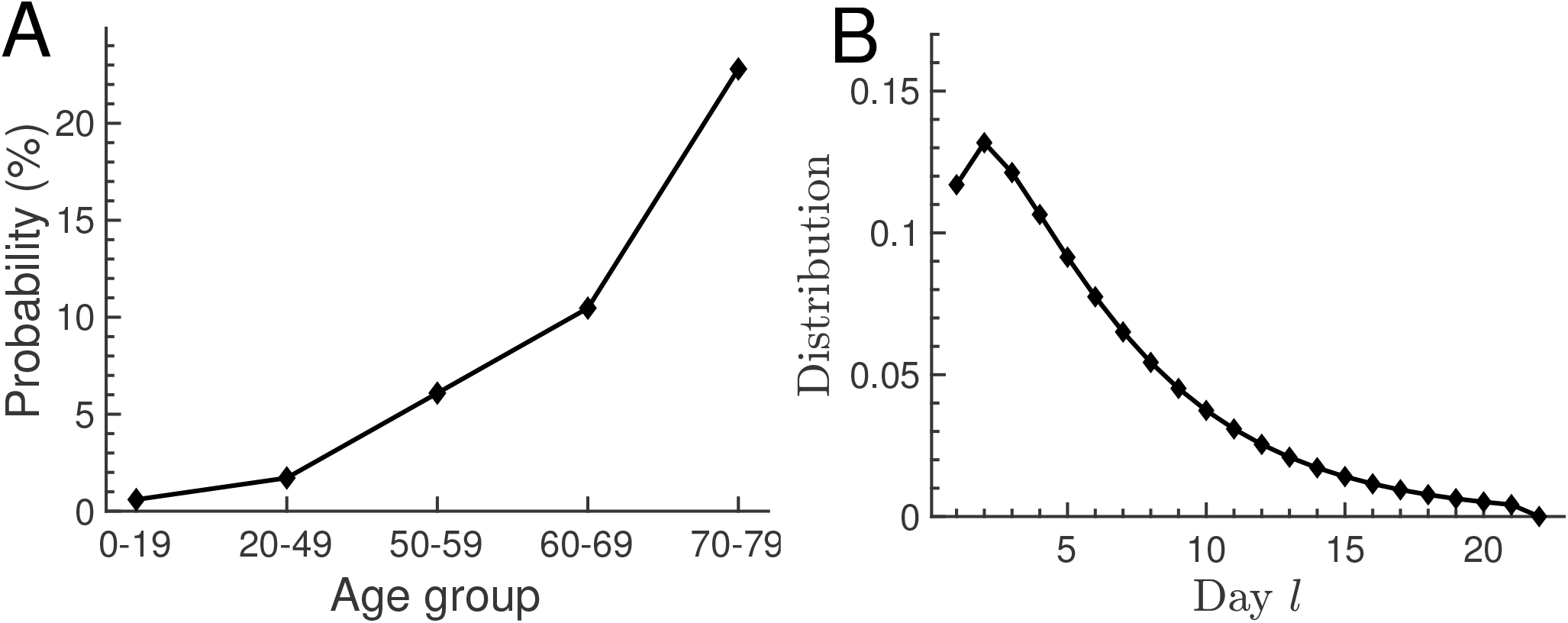
Probability and time distribution for a symptomatic person to become hospitalised. The mathematical expression for the fitting distribution is given by Eq. 26 with *l*_*max*_ = 22 days. (A) The fitting results for the age stratified probability that a symptomatic person becomes hospitalised are 0.6%, 1.7%, 6.1%, 10.5% and 22.8%. (B) Fitting result for the distribution that specifies at which day *l* after showing symptoms the hospitalisation occurs. We assumed the same distribution for all age groups. The mean number of days is 5.9.

**Figure S2:**
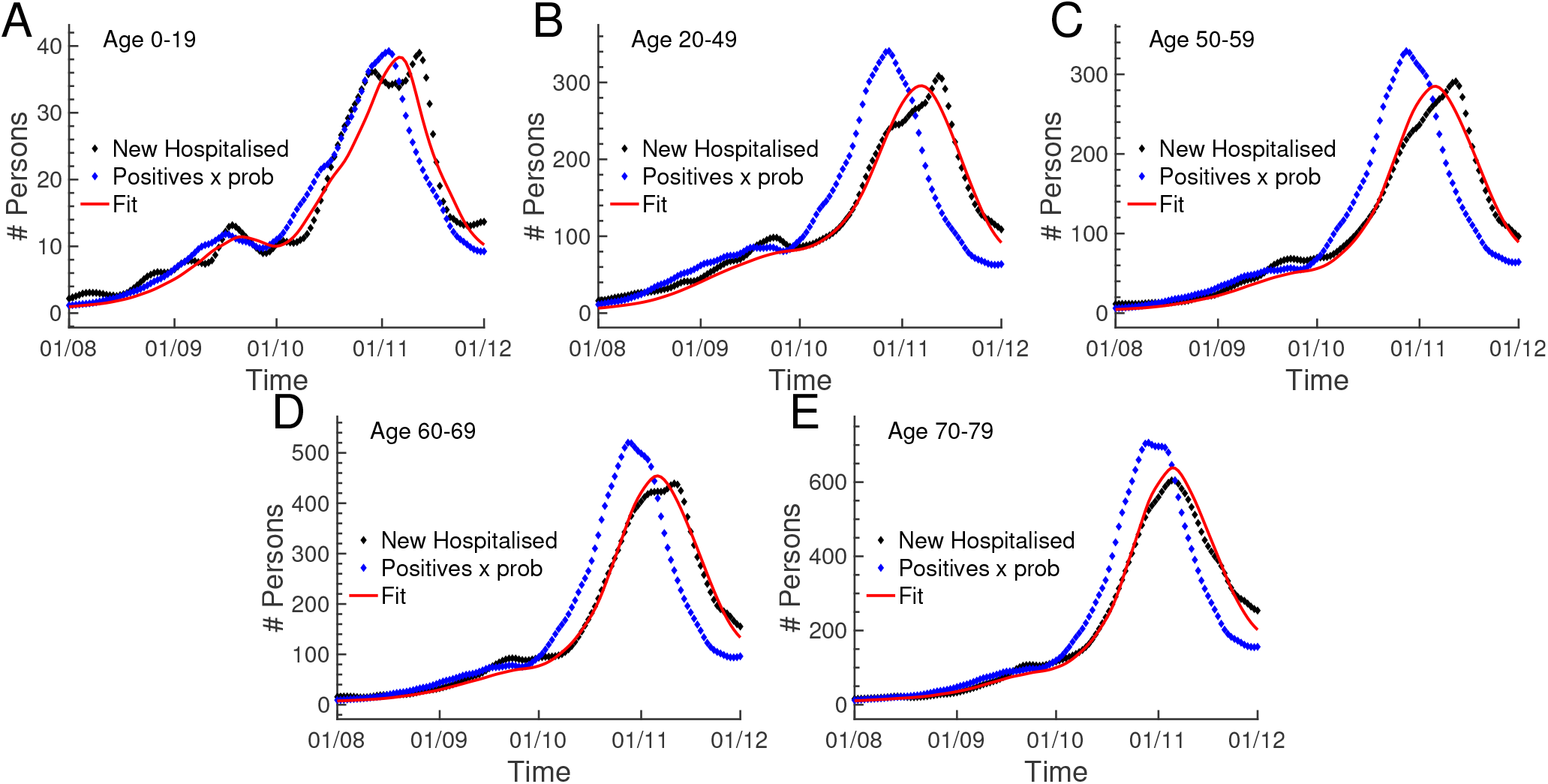
Correlation between positive testings and new hospitalisations. For each age group we fitted the probability that a positive tested person becomes hospitalised using the data for the number of new hospitalisations (black dots) and the data for the number of positive tested persons (blue dots; the data is multiplied by the fitted probability). The fitting result is shown in red. The mathematical expression for the fitting distribution is given by Eq. 26 with *l*_*max*_ = 21 days. The fitted probabilities are 0.6%, 1.5%, 4.5%, 10.5% and 22.8%.

### B Modeling framework

#### B.1 Model structure

The parameters, variables and probability distributions of the model are:

- *k*: Index for the age stratified groups.
- 1 ≤ *j* ≤ 5: Index of infection compartments.
- j=1 asymptomatic; j=2 symptomatic; j=3 in hospital; j=4 in ICU; j=5 in hospital after ICU.
- Time scales.
- *n*: Time measured as number of days since 01/01/2020.
- *l*: Number of days since an infected person joined its current compartment *j*.
- *h*: Vector that specifies the previous infection history. For example, the infection history of a hospitalised person that is currently in compartment *j* = 5 since *l* days comprises the number of days *l*_1_ that this person has been asymptomatic, the number of days *l*_2_ that it has been symptomatic, the number of hospitalisation days *l*_3_ before ICU, and the number of days *l*_4_ in ICU, such that *h* = (*l*_1_, *l*_2_, *l*_3_, *l*_4_). In this case the number of days since infection are *m* = *l*_1_ + *l*_2_ + *l*_3_ + *l*_4_ + *l*. The vector *h* is updated each time the infection compartment is changed. However, in the current implementation we consider only the time scales *n* and *l*.
- *S*_*k*_(*n*): Number of susceptible persons in group *k* at time *n*.
- Φ_*k*_(*n*) : Number of new infected in group *k* at time *n*.
- *I*_*k,h,j,l*_(*n*): Number of infected at time *n* belonging to age group *k*, infection history *h*, current compartment *j* at day *l* after joining this compartment. For *j* = 1 the infection history is fully determined by *l*, and here we can omit the index *h*.
- *p*_*k,h,j,l*;*j*_^*′*^ (*n*): Probability to transition from compartment *j* at day *l* to compartment *j*^*′*^ ≠ *j* for an infected in age group *k* with history *h*. The index *n* indicates that these probabilities might change over time due to social measures or modified hospital procedures.
- *p*_*k,h,j,l*;*rec*_(*n*): Probability to recover from compartment *j*.
- *p*_*k,h,j,l*;*dec*_(*n*): Probability to die in compartment *j*.
- *p*_*k,h,j,l*;*j*_ (*n*) = 1 − _*j*_^*′*^ _*≠j*_ *p*_*k,h,j,l*;*j*_^*′*^ (*n*) − *p*_*k,h,j,l*;*rec*_(*n*) − *p*_*k,h,j,l*;*dec*_(*n*): Probability to remain in compartment *j*.
- *R*_*k,h,j,l*_(*n*) = *p*_*k,h,j,l*;*rec*_(*n*)*I*_*k,h,j,l*_(*n*): Number of person that recover at time *n* from compartment *j*.
- *D*_*k,h,j,l*_(*n*) = *p*_*k,h,j,l*;*dec*_(*n*)*I*_*k,h,j,l*_(*n*): Number of persons that die in compartment *j* at time *n*. We consider that infected only die in ICU (*j* = 4).

The recurrence relations for the time evolution are

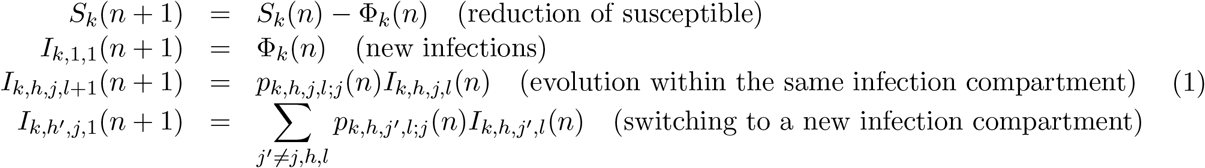

For each age group *k* we have the conservation equation

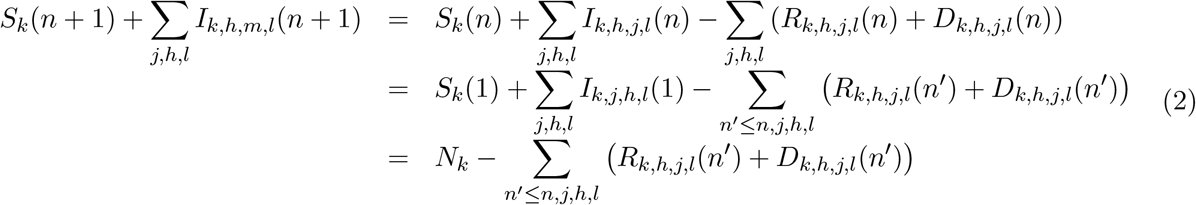

where *N*_*k*_ is the initial population. If *n* = 1 corresponds to the very beginning of the pandemic, we have *N*_*k*_ = *S*_*k*_(1) + Φ_*k*_(0), where Φ_*k*_(0) is the initial seed of new infections.

### B.2 New infections

We assume that the number of new infections Φ_*k*_(*n*) in group *k* at day *n* is proportional to the daily number of contacts C_*k*;*k*_^*′*^_,*j,m,l*_(*n*) that susceptible persons in group *k* make with infected persons from groups *k*^*′*^ in compartment *j*,

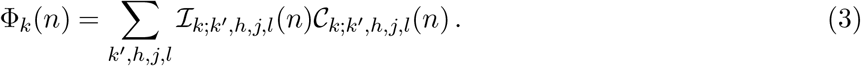

The proportionality constants are the infectivity parameters ℐ_*k*;*k*_^*′*^_,*h,j,l*_(*n*). We simplify and assume that as soon as the infection manifests itself by showing symptoms (switching to compartment *j* ≥ 2) an infected person takes precautions to avoid infecting others, in which case new infections only occur in interactions with asymptomatic persons (*j* = 1). With this assumption we have (note that *h* = *l* for *j* = 1, and we omit *h* in the following)

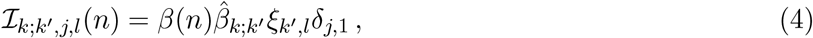

where *δ*_*i,j*_ is the Kronecker delta. The matrix 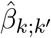models group specificities in the infection dynamics (we choose a normalization 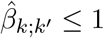; the matrix 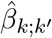 does not have to be symmetric). For example, with 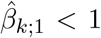and 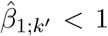 we consider the possibility that age group 1 is less infectious and less susceptible to infections. The parameters *ξ*_*k*_^*′*^,*l* describe how the contagiousness evolves as a function of the time *l* after infection (we normalize and use *ξ*_*k*_^*′*^_,*l*_ ≤ 1). The time dependent infectivity parameter *β*(*n*) accounts for changes that equally affect all groups, e.g. the introduction of masks. The initial value of *β*(*n*) depends on the normalisations of the other parameters. Moreover, if the affinity for mask wearing would be different among age groups, one would have to consider a time dependent matrix 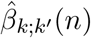.

To approximate the interactions between susceptible and asymptomatic persons we use

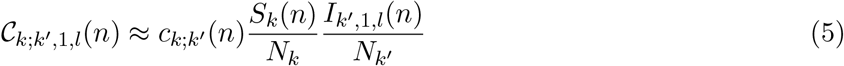

where *c*_*k*;*k*_^*′*^ (*n*) is the total daily number of contacts between groups *k* and *k*^*′*^, and 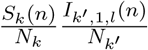 is the fraction of these contacts that correspond to encounters between susceptible and asymptomatic. The contact matrix *c*_*k*;_^*′*^ (*n*) is symmetric by definition, *c*_*k*;*k*_^*′*^ (*n*) = *c*_*k*_^*′*^_;*k*_ (*n*). The matrix is time dependent to account for social measures. Before lockdown we use [37]

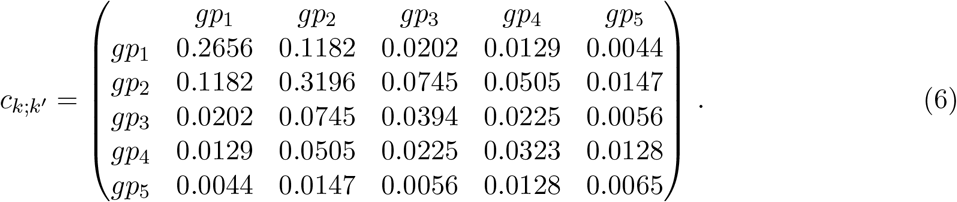

The contact matrix in Eq. 6 is normalized such that the total number of contacts is one, _*∑k,k*_^*′*^_*≥k*_ *c*_*k*;*k*_^*′*^ = 1.

Finally, with Eqs. 3–5 the number of new infections at day *n* generated in group *k* by infectious persons is

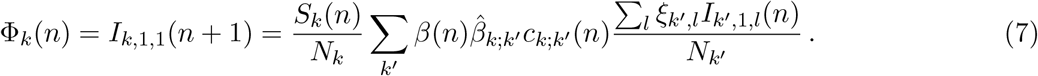

With

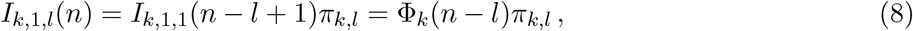

where *π*_*k,l*_ is the probability that an infected person is still asymptomatic after *l* days, and

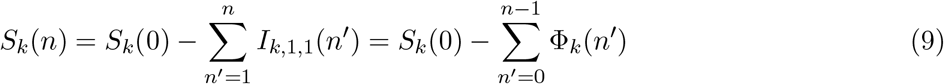

we obtain the recurrence relations

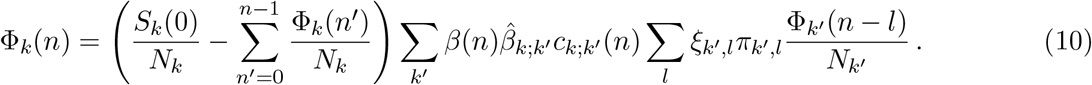

### B.3 Effective reproduction numbers

To connect to effective reproduction numbers (when the population is not any more totally susceptible), we consider the number of new infections that will be generated in age group *k* by a newly infected person in age group *k*^*′*^. By neglecting changes during the short contagious period, we obtain from Eq. 10 the reproduction matrix

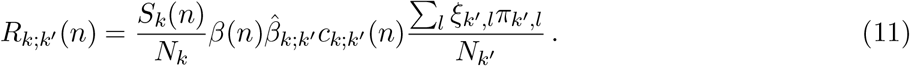

By considering the total number of new infections that will be generated by this infected person in group *k*^*′*^, we obtain the group dependent reproduction numbers

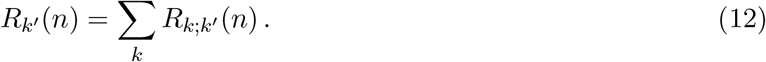

Finally, with the new infected persons Φ_*k*_(*n*) that are present at day *n*, we define the average reproduction number

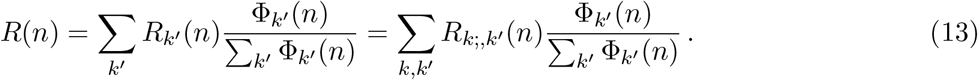

The pandemic grows for *R*(*n*) *>* 1 because the generated new infections ∑_*k*_^*′*^ *R*_*k*_^*′*^ (*n*)Φ_*k*_^*′*^ (*n*) is larger than the current value ∑_*k*_^*′*^ Φ_*k*_^*′*^ (*n*).

To characterize how the generated numbers of new infections will be distributed over the age groups, we define the reproduction distribution

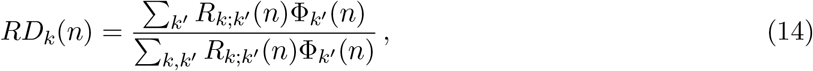

such that ∑_*k*_ *RD*_*k*_(*n*) = 1. To characterize whether the infection grows or declines in age group *k*, we define the reproduction growth index

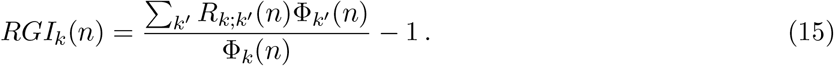

The pandemic grows in group *k* for *RGI*_*k*_(*n*) *>* 0.

### B.4 Transition probabilities

The model probabilities *p*_*k,h,j,l*;*i*_(*n*_0_) to transition from compartment *j* to compartment *i* (the label *i* also comprises the deceased and recovered compartment) are computed from the overall probabilities 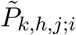 to transition from compartment *j* to compartment *i*, and the distributions 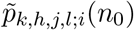 that specify at which day *l* this transition occurs. The time *n*_0_ indicates that these probabilities might change at time *n*_0_. We simplify the notation and omit the labels *k, h* and *n*_0_ for the following calculations.

The probabilities *p*_*j,l*;*i*_ satisfy the normalisation conditions ∑_*i*_ *p*_*j,l*;*i*_ = 1. In contrast, 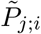 and 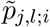 satisfy the normalization conditions 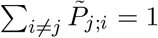 and 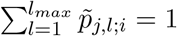. The probability 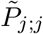 to remain in a compartment *j* is zero, since all infected will eventually recover or die. For numerical reasons, we truncate 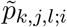 at a value *l*_*max*_ that is chosen sufficiently large. We chose *l*_*max*_ = 60 for *j* = 3, 4 and *l*_*max*_ = 80 for *j* = 5, since it takes longer for hospitalized to recover after ICU. As a function of 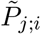 and 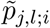 the probabilities *p*_*j,l*;*i*_ can be computed as

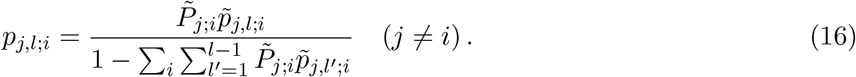

and *p*_*j,l*;*j*_ = 1 − ∑_*i*_ *≠ j p*_*j,l*;*i*_. Because the probability to be found in compartment *j* after *l*_*max*_ days is zero, we have 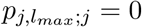.

### B.5 Equations to compute the number of infected in a compartment as a function of influx and initial condition

We now derive matrix equations to compute the time evolution of the number of infected in a compartment *j* with a given initial condition and a prescribed influx. We need these expression to implement fitting procedures. The probability to be found in compartment *j* after *l* days is

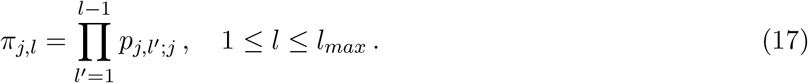

The occupancy of compartment *j* at time *n* consists of persons that originate from an initial condition, and persons that joined this compartment via influx from other compartments.

#### B.5.1 0ccupancy and fluxes to other compartments as a function of a given influx

The occupancy *O*_*j*_(*n*) due to the influx *I*_*j*,1_(*n*) from other compartments is given by the convolution

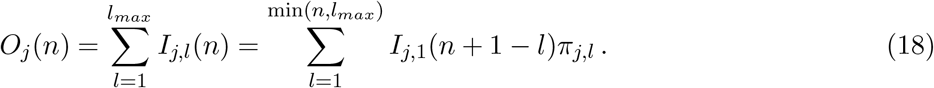

The number of persons *C*_*j*;*i*_(*n*) that change to a different compartment *i* is

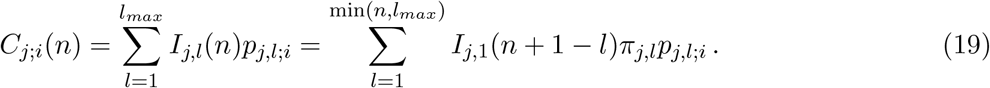

The number of new persons in compartment *i* that originate from compartment *j* is *I*_*i*,1;*j*_(*n* + 1) = *C*_*j*;*i*_(*n*).

#### B.5.2 0ccupancy and fluxes to other compartments as a function of an initial condition

We now consider the occupancy that originates from an initial condition *I*_*j,l*_(*n*_0_) at day *n*_0_ (without loss of generality we consider *n*_0_ = 1). The number of infected at a future day *n* that are still in compartment *j* is

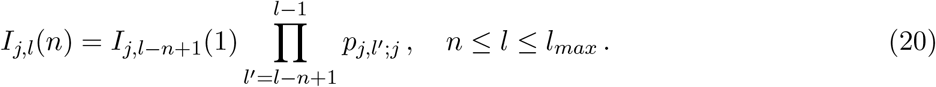

The occupancy 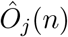 due to the initial condition therefore is

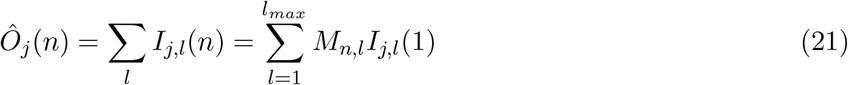

Where

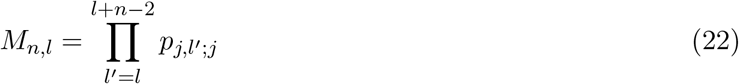

For *n* + *l > l*_*max*_ + 2 we define *M*_*n,l*_ = 0.

The number of persons that change to a different compartment *i* is

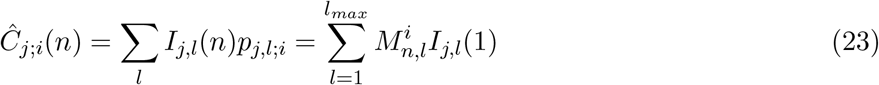

With

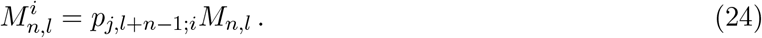

For *l* + *n > l*_*max*_ + 1 we define 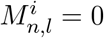.

### B.6 Method to implement parameter changes

To implement parameter changes from *P*_*old*_ to *P*_*new*_ at day *n*_0_ we use

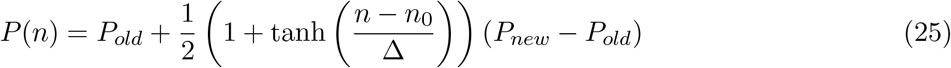

with Δ = 1.

### B.7 Discrete Gamma distribution

To fit distributions we use the discrete Gamma distribution defined as [61]

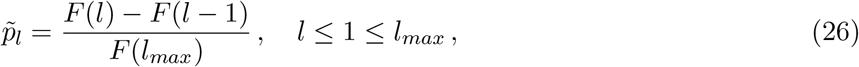

with

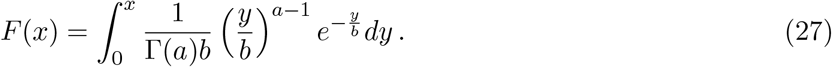

The distribution depends on two parameters *a >* 0 and *b >* 0.

### B.8 Fitting procedure

For the fitting procedure we used the constrained optimization procedure based on MATLAB’s *fminsearch* procedure [62]. We use the matrix equations derived section B.5 to sequentially compute for each compartment the occupancy and outflux to other compartments as function of the initial condition and the influx from other compartments. For example, with the computed influx of new hospitalisations, we compute the future occupancies and fluxes between hospital compartments (as well as number of death and recovered) as a function the switching parameters. We then use data, simulation results and a cost function (here we chose the mean squared error) together with the constrained optimization procedure to fit the specified parameters.

## Declarations

## Author contributions

JR and DH designed research. JR designed the model, performed analysis, developed fitting procedures and implemented the model in MATLAB. SR and JFT provided clinical data. AP collected, processed and analysed data. JR and DH wrote the manuscript.

## Code availability

The code is available upon request to the authors.

## Data availability

The data is available upon request to the authors.

## Ethics approval

The clinical data comes from the OutcomeRea database that was declared to the *Commission Nationale de l’Informatique et des Libertés* (#999,262), in accordance with French law, and this study was approved by the institutional review board of Clermont Ferrand. I

## Consent to participate

Consent is not required because the study did not modify patients management and the data were anonymously collected.

## Acknowledgements

We thank Dr. Dan Longrois for discussions.

## References

[1] S. Flaxman, S. Mishra, A. Gandy, H. J. T. Unwin, T. A. Mellan, H. Coupland, C. Whittaker, H. Zhu, T. Berah, J. W. Eaton, M. Monod, P. N. Perez-Guzman, N. Schmit, L. Cilloni, K. E. C. Ainslie, M. Baguelin, A. Boonyasiri, O. Boyd, L. Cattarino, L. V. Cooper, Z. Cucunubá, G. Cuomo-Dannenburg, A. Dighe, B. Djaafara, I. Dorigatti, S. L. van Elsland, R. G. FitzJohn, K. A. M. Gaythorpe, L. Geidelberg, N. C. Grassly, W. D. Green, T. Hallett, A. Hamlet, W. Hinsley, B. Jeffrey, E. Knock, D. J. Laydon, G. Nedjati-Gilani, P. Nouvellet, K. V. Parag, I. Siveroni, H. A. Thompson, R. Verity, E. Volz, C. E. Walters, H. Wang, Y. Wang, O. J. Watson, P. Winskill, X. Xi, P. G. T. Walker, A. C. Ghani, C. A. Donnelly, S. Riley, M. A. C. Vollmer, N. M. Ferguson, L. C. Okell, S. Bhatt, and I. C. C.-. R. Team, “Estimating the effects of non-pharmaceutical interventions on covid-19 in europe,” Nature, vol. 584, no. 7820, pp. 257–261, 2020.

[2] H. Salje, C. Tran Kiem, N. Lefrancq, N. Courtejoie, P. Bosetti, J. Paireau, A. Andronico, N. Hozé, J. Richet, C.-L. Dubost, Y. Le Strat, J. Lessler, D. Levy-Bruhl, A. Fontanet, L. Opatowski, P.-Y. Boelle, and S. Cauchemez, “Estimating the burden of sars-cov-2 in france,” Science, vol. 369, no. 6500, pp. 208–211, 2020.

[3] L. Di Domenico, G. Pullano, C. E. Sabbatini, P.-Y. Boëlle, and V. Colizza, “Impact of lockdown on covid-19 epidemic in île-de-france and possible exit strategies,” BMC Medicine, vol. 18, no. 1, p. 240, 2020.

[4] K. Leung, J. T. Wu, D. Liu, and G. M. Leung, “First-wave covid-19 transmissibility and severity in china outside hubei after control measures, and second-wave scenario planning: a modelling impact assessment,” The Lancet, vol. 395, no. 10233, pp. 1382–1393, 2020.

[5] T. N. Starr, A. J. Greaney, A. Addetia, W. W. Hannon, M. C. Choudhary, A. S. Dingens, J. Z. Li, and J. D. Bloom, “Prospective mapping of viral mutations that escape antibodies used to treat covid-19,” Science, vol. 371, no. 6531, pp. 850–854, 2021.

[6] R. N. Thompson, “Epidemiological models are important tools for guiding covid-19 interventions,” BMC medicine, vol. 18, no. 1, pp. 1–4, 2020.

[7] M. Nicola, Z. Alsafi, C. Sohrabi, A. Kerwan, A. Al-Jabir, C. Iosifidis, M. Agha, and R. Agha, “The socio-economic implications of the coronavirus pandemic (covid-19): A review,” International Journal of Surgery, vol. 78, pp. 185–193, 2020.

[8] R. N. Thompson, T. D. Hollingsworth, V. Isham, D. Arribas-Bel, B. Ashby, T. Britton, P. Challenor, L. H. Chappell, H. Clapham, N. J. Cunniffe, et al., “Key questions for modelling covid-19 exit strategies,” Proceedings of the Royal Society B, vol. 287, no. 1932, p. 20201405, 2020.

[9] J. Dehning, J. Zierenberg, F. P. Spitzner, M. Wibral, J. P. Neto, M. Wilczek, and V. Priesemann, “Inferring change points in the spread of covid-19 reveals the effectiveness of interventions,” Science, vol. 369, no. 6500, 2020.

[10] J. T. Wu, K. Leung, and G. M. Leung, “Nowcasting and forecasting the potential domestic and international spread of the 2019-ncov outbreak originating in wuhan, china: a modelling study,” The Lancet, vol. 395, pp. 689–697, 2021/04/14 2020.

[11] R. C. Reiner, R. M. Barber, J. K. Collins, P. Zheng, C. Adolph, J. Albright, C. M. Antony, A. Y. Aravkin, S. D. Bachmeier, B. Bang-Jensen, M. S. Bannick, S. Bloom, A. Carter, E. Castro, K. Causey, S. Chakrabarti, F. J. Charlson, R. M. Cogen, E. Combs, X. Dai, W. J. Dangel, L. Earl, S. B. Ewald, M. Ezalarab, A. J. Ferrari, A. Flaxman, J. J. Frostad, N. Fullman, E. Gakidou, J. Gallagher, S. D. Glenn, E. A. Goosmann, J. He, N. J. Henry, E. N. Hulland, B. Hurst, C. Johanns, P. J. Kendrick, A. Khemani, S. L. Larson, A. Lazzar-Atwood, K. E. LeGrand, H. Lescinsky, A. Lindstrom, E. Linebarger, R. Lozano, R. Ma, J. Mansson, B. Magistro, A. M. M. Herrera, L. B. Marczak, M. K. Miller-Petrie, A. H. Mokdad, J. D. Morgan, P. Naik, C. M. Odell, J. K. O’Halloran, A. E. Osgood-Zimmerman, S. M. Ostroff, M. Pasovic, L. Penberthy, G. Phipps, D. M. Pigott, I. Pollock, R. E. Ramshaw, S. B. Redford, G. Reinke, S. Rolfe, D. F. Santomauro, J. R. Shackleton, D. H. Shaw, B. S. Sheena, A. Sholokhov, R. J. D. Sorensen, G. Sparks, E. E. Spurlock, M. L. Subart, R. Syailendrawati, A. E. Torre, C. E. Troeger, T. Vos, A. Watson, S. Watson, K. E. Wiens, L. Woyczynski, L. Xu, J. Zhang, S. I. Hay, S. S. Lim, C. J. L. Murray, and I. H. C.-. F. Team, “Modeling covid-19 scenarios for the united states,” Nature Medicine, vol. 27, no. 1, pp. 94–105, 2021.

[12] T. Britton, F. Ball, and P. Trapman, “A mathematical model reveals the influence of population heterogeneity on herd immunity to sars-cov-2,” Science, vol. 369, no. 6505, pp. 846–849, 2020.

[13] Q. Griette and P. Magal, “Clarifying predictions for covid-19 from testing data: The example of new york state,” Infectious Disease Modelling, vol. 6, pp. 273–283, 2021.

[14] Z. Liu, P. Magal, and G. Webb, “Predicting the number of reported and unreported cases for the covid-19 epidemics in china, south korea, italy, france, germany and united kingdom,” Journal of theoretical biology, vol. 509, p. 110501, 2021.

[15] Z. Liu, P. Magal, O. Seydi, and G. Webb, “Understanding unreported cases in the covid-19 epidemic outbreak in wuhan, china, and the importance of major public health interventions,” Biology, vol. 9, no. 3, p. 50, 2020.

[16] S. Moore, E. M. Hill, M. J. Tildesley, L. Dyson, and M. J. Keeling, “Vaccination and nonpharmaceutical interventions for covid-19: a mathematical modelling study,” The Lancet Infectious Diseases, vol. 21, no. 6, pp. 793–802, 2021.

[17] V. Volpert, M. Banerjee, and S. Petrovskii, “On a quarantine model of coronavirus infection and data analysis,” Mathematical Modelling of Natural Phenomena, vol. 15, p. 24, 2020.

[18] M. J. Keeling, E. Hill, E. Gorsich, B. Penman, G. Guyver-Fletcher, A. Holmes, T. Leng, H. McKimm, M. Tamborrino, L. Dyson, and M. Tildesley, “Predictions of covid-19 dynamics in the uk: short-term forecasting and analysis of potential exit strategies,” PLOS Computational Biology, vol. 17, pp. 1–20, 01 2021.

[19] R. Verity, L. C. Okell, I. Dorigatti, P. Winskill, C. Whittaker, N. Imai, G. Cuomo-Dannenburg, H. Thompson, P. G. T. Walker, H. Fu, A. Dighe, J. T. Griffin, M. Baguelin, S. Bhatia, A. Boonyasiri, A. Cori, Z. Cucunubá, R. FitzJohn, K. Gaythorpe, W. Green, A. Hamlet, W. Hinsley, D. Laydon, G. Nedjati-Gilani, S. Riley, S. van Elsland, E. Volz, H. Wang, Y. Wang, X. Xi, C. A. Donnelly, A. C. Ghani, and N. M. Ferguson, “Estimates of the severity of coronavirus disease 2019: a model-based analysis,” The Lancet Infectious Diseases, vol. 20, no. 6, pp. 669–677, 2020.

[20] A. J. Kucharski, T. W. Russell, C. Diamond, Y. Liu, J. Edmunds, S. Funk, R. M. Eggo, F. Sun, M. Jit, J. D. Munday, N. Davies, A. Gimma, K. van Zandvoort, H. Gibbs, J. Hellewell, C. I. Jarvis, S. Clifford, B. J. Quilty, N. I. Bosse, S. Abbott, P. Klepac, and S. Flasche, “Early dynamics of transmission and control of covid-19: a mathematical modelling study,” The Lancet Infectious Diseases, vol. 20, no. 5, pp. 553–558, 2020.

[21] L. Roques, E. K. Klein, J. Papaïx, A. Sar, and S. Soubeyrand, “Impact of lockdown on the epidemic dynamics of covid-19 in france,” Frontiers in medicine, vol. 7, p. 274, 2020.

[22] L. Roques, E. K. Klein, J. Papaix, A. Sar, and S. Soubeyrand, “Using early data to estimate the actual infection fatality ratio from covid-19 in france,” Biology, vol. 9, no. 5, p. 97, 2020.

[23] M. Gatto, E. Bertuzzo, L. Mari, S. Miccoli, L. Carraro, R. Casagrandi, and A. Rinaldo, “Spread and dynamics of the covid-19 epidemic in italy: Effects of emergency containment measures,” Proceedings of the National Academy of Sciences, vol. 117, no. 19, pp. 10484–10491, 2020.

[24] L. Mari, R. Casagrandi, E. Bertuzzo, D. Pasetto, S. Miccoli, A. Rinaldo, and M. Gatto, “The epidemicity index of recurrent sars-cov-2 infections,” Nature communications, vol. 12, no. 1, pp. 1–12, 2021.

[25] E. Bertuzzo, L. Mari, D. Pasetto, S. Miccoli, R. Casagrandi, M. Gatto, and A. Rinaldo, “The geography of covid-19 spread in italy and implications for the relaxation of confinement measures,” Nature communications, vol. 11, no. 1, pp. 1–11, 2020.

[26] L. Di Domenico, G. Pullano, C. E. Sabbatini, P.-Y. Boëlle, and V. Colizza, “Modelling safe protocols for reopening schools during the covid-19 pandemic in france,” Nature communications, vol. 12, no. 1, pp. 1–10, 2021.

[27] G. Pullano, L. Di Domenico, C. E. Sabbatini, E. Valdano, C. Turbelin, M. Debin, C. Guerrisi, C. Kengne-Kuetche, C. Souty, T. Hanslik, T. Blanchon, P.-Y. Boëlle, J. Figoni, S. Vaux, C. Campèse, S. Bernard-Stoecklin, and V. Colizza, “Underdetection of cases of covid-19 in france threatens epidemic control,” Nature, vol. 590, no. 7844, pp. 134–139, 2021.

[28] F. Pinotti, L. Di Domenico, E. Ortega, M. Mancastroppa, G. Pullano, E. Valdano, P.-Y. Boëlle, C. Poletto, and V. Colizza, “Tracing and analysis of 288 early sars-cov-2 infections outside china: A modeling study,” PLOS Medicine, vol. 17, no. 7, pp. 1–13, 2020.

[29] N. Hoertel, M. Blachier, C. Blanco, M. Olfson, M. Massetti, M. S. Rico, F. Limosin, and H. Leleu, “A stochastic agent-based model of the sars-cov-2 epidemic in france,” Nature medicine, vol. 26, no. 9, pp. 1417–1421, 2020.

[30] C. Selinger, M. Choisy, and S. Alizon, “Predicting covid-19 incidence in french hospitals using human contact network analytics,” International Journal of Infectious Diseases, vol. 111, pp. 100–107, 2021.

[31] F. Balabdaoui and D. Mohr, “Age-stratified discrete compartment model of the covid-19 epidemic with application to switzerland,” Scientific Reports, vol. 10, no. 1, p. 21306, 2020.

[32] J. Reingruber, A. Papale, and D. Holcman, “Monitoring and predicting sars-cov-2 epidemic in france after deconfinement using a multiscale and age-dependent model,” medRxiv, 2020.

[33] P. Bosetti, C. T. Kiem, Y. Yazdanpanah, A. Fontanet, B. Lina, V. Colizza, and S. Cauchemez, “Impact of mass testing during an epidemic rebound of sars-cov-2: a modelling study using the example of france,” Eurosurveillance, vol. 26, no. 1, 2021.

[34] Q. Richard, S. Alizon, M. Choisy, M. T. Sofonea, and R. Djidjou-Demasse, “Age-structured non-pharmaceutical interventions for optimal control of covid-19 epidemic,” PLoS computational biology, vol. 17, no. 3, p. e1008776, 2021.

[35] “https://geodes.santepubliquefrance.fr.”

[36] “https://www.data.gouv.fr/fr/datasets/donnees-hospitalieres-relatives-a-lepidemie-de-covid-19.”

[37] G. Béraud, S. Kazmercziak, P. Beutels, D. Levy-Bruhl, X. Lenne, N. Mielcarek, Y. Yazdanpanah, P.-Y. Boëlle, N. Hens, and B. Dervaux, “The french connection: The first large population-based contact survey in france relevant for the spread of infectious diseases,” PLOS ONE, vol. 10, no. 7, pp. 1–22, 2015.

[38] S. A. Lauer, K. H. Grantz, Q. Bi, F. K. Jones, Q. Zheng, H. R. Meredith, A. S. Azman, N. G. Reich, and J. Lessler, “The incubation period of coronavirus disease 2019 (covid-19) from publicly reported confirmed cases: Estimation and application.,” Annals of Internal Medicine, vol. 172, no. 9, pp. 577–582, 2020.

[39] E. Lavezzo, E. Franchin, C. Ciavarella, G. Cuomo-Dannenburg, L. Barzon, C. Del Vecchio, L. Rossi, R. Manganelli, A. Loregian, N. Navarin, D. Abate, M. Sciro, S. Merigliano, E. De Canale, M. C. Vanuzzo, V. Besutti, F. Saluzzo, F. Onelia, M. Pacenti, S. G. Parisi, G. Carretta, D. Donato, L. Flor, S. Cocchio, G. Masi, A. Sperduti, L. Cattarino, R. Salvador, M. Nicoletti, F. Caldart, G. Castelli, E. Nieddu, B. Labella, L. Fava, M. Drigo, K. A. M. Gaythorpe, A. R. Brazzale, S. Toppo, M. Trevisan, V. Baldo, C. A. Donnelly, N. M. Ferguson, I. Dorigatti, A. Crisanti, K. E. C. Ainslie, M. Baguelin, S. Bhatt, A. Boonyasiri, O. Boyd, H. L. Coupland, Z. Cucunubá, B. A. Djafaara, C. A. Donnelly, S. L. van Elsland, R. FitzJohn, S. Flaxman, K. A. M. Gaythorpe, W. D. Green, T. Hallett, A. Hamlet, D. Haw, N. Imai, B. Jeffrey, E. Knock, D. J. Laydon, T. Mellan, S. Mishra, G. Nedjati-Gilani, P. Nouvellet, L. C. Okell, K. V. Parag, S. Riley, H. A. Thompson, H. J. T. Unwin, R. Verity, M. A. C. Vollmer, P. G. T. Walker, C. E. Walters, H. Wang, Y. Wang, O. J. Watson, C. Whittaker, L. K. Whittles, X. Xi, N. M. Ferguson, and I. C. C.-. R. Team, “Suppression of a sars-cov-2 outbreak in the italian municipality of vo’,” Nature, vol. 584, no. 7821, pp. 425–429, 2020.

[40] T. C. Jones, B. Mühlemann, T. Veith, G. Biele, M. Zuchowski, J. Hofmann, A. Stein, A. Edelmann, V. M. Corman, and C. Drosten, “An analysis of sars-cov-2 viral load by patient age,” medRxiv, 2020.

[41] D. Cereda, M. Tirani, F. Rovida, V. Demicheli, M. Ajelli, P. Poletti, F. Trentini, G. Guzzetta, V. Marziano, A. Barone, M. Magoni, S. Deandrea, G. Diurno, M. Lombardo, M. Faccini, A. Pan, R. Bruno, E. Pariani, G. Grasselli, A. Piatti, M. Gramegna, F. Baldanti, A. Melegaro, and S. Merler, “The early phase of the covid-19 outbreak in lombardy, italy,” arxive, 2020.

[42] A. Fontanet, R. Grant, L. Tondeur, Y. Madec, L. Grzelak, I. Cailleau, M.-N. Ungeheuer, C. Renaudat, S. F. Pellerin, L. Kuhmel, et al., “Sars-cov-2 infection in primary schools in northern france: A retrospective cohort study in an area of high transmission,” MedRxiv, 2020.

[43] L. Di Domenico, G. Pullano, C. E. Sabbatini, Boëlle, Pierre-Yves, and V. Colizza, “Report 9, expected impact of lockdown in île-de-france and possible exit strategies,” 2020.

[44] K. K. Cheng, T. H. Lam, and C. C. Leung, “Wearing face masks in the community during the covid-19 pandemic: altruism and solidarity,” The Lancet, 2020.

[45] N. Hozé, J. Paireau, N. Lapidus, C. T. Kiem, H. Salje, G. Severi, M. Touvier, M. Zins, X. de Lamballerie, D. Lévy-Bruhl, et al., “Monitoring the proportion of the population infected by sars-cov-2 using age-stratified hospitalisation and serological data: a modelling study,” The Lancet Public Health, 2021.

[46] R. Li, S. Pei, B. Chen, Y. Song, T. Zhang, W. Yang, and J. Shaman, “Substantial undocumented infection facilitates the rapid dissemination of novel coronavirus (sars-cov-2),” Science, vol. 368, no. 6490, pp. 489–493, 2020.

[47] A. T. Levin, W. P. Hanage, N. Owusu-Boaitey, K. B. Cochran, S. P. Walsh, and G. Meyerowitz-Katz, “Assessing the age specificity of infection fatality rates for covid-19: systematic review, meta-analysis, and public policy implications,” European Journal of Epidemiology, vol. 35, no. 12, pp. 1123–1138, 2020.

[48] S. M. Moghadas, M. C. Fitzpatrick, P. Sah, A. Pandey, A. Shoukat, B. H. Singer, and A. P. Galvani, “The implications of silent transmission for the control of covid-19 outbreaks,” Proceedings of the National Academy of Sciences, vol. 117, no. 30, pp. 17513–17515, 2020.

[49] K. Mizumoto, K. Kagaya, A. Zarebski, and G. Chowell, “Estimating the asymptomatic proportion of coronavirus disease 2019 (covid-19) cases on board the diamond princess cruise ship, yokohama, japan,” Euro Surveill, vol. 25, no. 10, 2020.

[50] H. Streeck, B. Schulte, B. M. Kümmerer, E. Richter, T. Höller, C. Fuhrmann, E. Bartok, R. Dolscheid-Pommerich, M. Berger, L. Wessendorf, M. Eschbach-Bludau, A. Kellings, A. Schwaiger, M. Coenen, P. Hoffmann, B. Stoffel-Wagner, M. M. Nöthen, A. M. Eis-Hübinger, M. Exner, R. M. Schmithausen, M. Schmid, and G. Hartmann, “Infection fatality rate of sars-cov2 in a super-spreading event in germany,” Nature Communications, vol. 11, no. 1, p. 5829, 2020.

[51] P. Zimmermann and N. Curtis, “Coronavirus infections in children including covid-19: an overview of the epidemiology, clinical features, diagnosis, treatment and prevention options in children,” The Pediatric infectious disease journal, vol. 39, no. 5, p. 355, 2020.

[52] N. G. Davies, P. Klepac, Y. Liu, K. Prem, M. Jit, and R. M. Eggo, “Age-dependent effects in the transmission and control of covid-19 epidemics,” Nature medicine, vol. 26, no. 8, pp. 1205–1211, 2020.

[53] C. Jiehao, X. Jin, L. Daojiong, Y. Zhi, X. Lei, Q. Zhenghai, Z. Yuehua, Z. Hua, J. Ran, L. Pengcheng, et al., “A case series of children with 2019 novel coronavirus infection: clinical and epidemiological features,” Clinical Infectious Diseases, vol. 71, no. 6, pp. 1547–1551, 2020.

[54] C. Chen, C. Zhu, D. Yan, H. Liu, D. Li, Y. Zhou, X. Fu, J. Wu, C. Ding, G. Tian, et al., “The epidemiological and radiographical characteristics of asymptomatic infections with the novel coronavirus (covid-19): A systematic review and meta-analysis,” International Journal of Infectious Diseases, 2021.

[55] L. A. Nikolai, C. G. Meyer, P. G. Kremsner, and T. P. Velavan, “Asymptomatic sars coronavirus 2 infection: Invisible yet invincible,” International Journal of Infectious Diseases, 2020.

[56] R. M. Viner, O. T. Mytton, C. Bonell, G. Melendez-Torres, J. Ward, L. Hudson, C. Waddington, J. Thomas, S. Russell, F. Van Der Klis, et al., “Susceptibility to sars-cov-2 infection among children and adolescents compared with adults: a systematic review and meta-analysis,” JAMA pediatrics, vol. 175, no. 2, pp. 143–156, 2021.

[57] J. Zhang, M. Litvinova, Y. Liang, Y. Wang, W. Wang, S. Zhao, Q. Wu, S. Merler, C. Viboud, A. Vespignani, et al., “Changes in contact patterns shape the dynamics of the covid-19 outbreak in china,” Science, vol. 368, no. 6498, pp. 1481–1486, 2020.

[58] M. ODriscoll, G. R. Dos Santos, L. Wang, D. A. Cummings, A. S. Azman, J. Paireau Fontanet, S. Cauchemez, and H. Salje, “Age-specific mortality and immunity patterns of sars-cov-2,” Nature, vol. 590, no. 7844, pp. 140–145, 2021.

[59] J. P. Ioannidis, “Reconciling estimates of global spread and infection fatality rates of covid-19: An overview of systematic evaluations,” European journal of clinical investigation, vol. 51, no. 5, p. e13554, 2021.

[60] S. Riley, K. Eames, V. Isham, D. Mollison, and P. Trapman, “Five challenges for spatial epidemic models,” Epidemics, vol. 10, pp. 68–71, 2015.

[61] Chakraborty, “Discrete gamma distributions: Properties and parameter estimations,” Communications in Statistics - Theory and Methods, vol. 41, no. 18, pp. 3301–3324, 2012.

[62] J. D’Errico, “fminsearchbnd, fminsearchcon (https://www.mathworks.com/matlabcentral/fileexchange/8277-fminsearchbnd-fminsearchcon),” MATLAB Central File Exchange, 2021.

